# Timing the Target: Exploring the Circadian Transcriptome of Biologic Therapies in Asthma

**DOI:** 10.64898/2026.11.16.26368198

**Authors:** Ayobami Akenroye, Vincent J. Carey, Frank A.J.L. Scheer, Scott T. Weiss

## Abstract

**Background:** Many of the immune pathways involved in asthma or in asthma treatment show circadian or diurnal variation. Understanding this variation can aid in biomarker discovery and/or therapy optimization.

**Objective:** To evaluate if receptors, cytokines, and their pathways (n=46) targeted by currently approved biologics show significant diurnal rhythmicity and to identify their peak expression time.

**Methods:** We used longitudinal bulk RNA sequencing datasets (discovery/replication cohorts: GSE113883 and GSE223613) and one microarray file (GSE39445) for cross-platform replication/validation. First, we used 16 canonical clock genes (including *CLOCK*, *BMAL1*, *REV-ERBß*) as positive controls to explore rhythmicity in each cohort. Thereafter, we evaluated if any targets of currently approved biologics or their corresponding receptors showed significant diurnal variation. For significant genes (Benjamini-Hochberg FDR<0.05), we estimated the baseline average, phase, peak-time of expression across the 24-hour day using clock time and quantified the cross-cohort consistency in the peak time of expression across the cohorts.

**Results:** These cohorts included 453 longitudinal samples collected every two to four hours over a 24-28h cycle. Twelve (75%) of 16 evaluable clock genes showed significant rhythmicity. Only 34 of 46 (74%) asthma-related genes were evaluable across two or more cohorts. Of these 34 genes, 23 (68%) showed significant variation. Across all cohorts, the most preserved rhythmically expressed targets included many pathways or targets of monoclonal antibodies that are currently approved or in development: *GATA3* and *STAT6*, master regulators of type 2 inflammation and IL4/IL13 signaling; chemokines, including the neutrophil chemoattractant, *CXCL8*; *IL-5RA*, benralizumab’s target; *FcRE1*, suppressed by omalizumab; and *IL1RL1*(*ST2*) in clinical trials.

**Conclusion:** Many therapeutic targets in asthma show significant diurnal rhythmicity. Future studies on how dosing time affects their therapeutic effectiveness while minimizing side effects are needed.

**Take Home Message:**

- Many targets of biologics and their pathways are significantly rhythmic in expression across the day. This includes *IL5RA*, transcription factors, like *STAT6* and *GATA3*, and the high-affinity IgE receptor. Asthma may be a prime area for chronotherapy.

**Highlights:**

- Longitudinal transcriptomics data show that many targets of currently available biologics and their pathways are significantly rhythmic in expression across the day.
- The circadian transcriptome includes surface receptors like *IL5RA*, targeted by benralizumab, and transcription factors, like *STAT6* and *GATA3*, master regulators of Th2 differentiation involved in the IL-4/-13 pathway targeted by dupilumab, and the high affinity IgE receptor suppressed by omalizumab.
- Monoclonal antibody therapy in asthma may be a prime area for investigating chronotherapy in asthma.

## INTRODUCTION

The cyclical nature of asthma symptoms have been known since antiquity, with most patients with persistent asthma exhibiting features of ‘nocturnal asthma’.^1–4^ Asthma symptoms peak at night and associated factors such as allergic rhinitis and upper respiratory tract infection symptoms are often preferentially worse at night. Relatedly, a nighttime dip in peak expiratory flow, or an increase in nighttime peak flow variability, is often the first sign of an impending asthma flare.^2,5^ Furthermore, biomarkers commonly used in defining asthma phenotypes, including the peripheral blood eosinophil count and exhaled nitric oxide, oscillate markedly over the day, with higher values at night or in the early morning respectively.^6–8^ Additionally, glucocorticoids, the mainstay of asthma treatment, and many of the immune pathways involved in asthma pathogenesis, show significant diurnal and/or circadian rhythmicity.^3,5,9^ Patients with more severe asthma and relatively worse lung function often have the greatest fluctuations in nighttime to daytime symptoms, suggesting that these diurnal patterns may be associated with intrinsic asthma severity and/or control.^2^ There is strong evidence that such diurnal variations in patients with asthma have an important underlying circadian mechanism, i.e., independent of effects of the sleep-wake, rest-activity, fasting-eating, dark-light, and environmental temperature cycles.^2,10^

Over forty percent of the mammalian genome is under circadian clock control, with the liver, kidney, and lungs having the highest proportion of protein-encoding genes with demonstrable circadian rhythmicity in rodents.^11^ The central clock, which includes the *BMAL1*-*CLOCK* heterodimer, regulates a wide range of cellular functions, including mast cell activation, monocyte chemoattraction, anti-bacterial and anti-viral immunity, and tissue repair.^12–14^ Although mammalian organs, like the lungs, have their own intrinsic circadian rhythm, the core clock genes often oscillate synchronously across organs and manipulation of clock genes can lead to pro- or anti-inflammatory effects.^11,12^ In addition to this central rhythm, multiple environmental factors such as light exposure, hormones, including catecholamines and glucocorticoids, exercise, food intake and timing of food intake, and temperature can all impact this circadian rhythm. Multiple medications show a time-of-day effect and many medications impact clock-controlled genes (CCG) or genes with significant circadian rhythmicity.^11,15^ There is emerging evidence that there are time-of-day effects to omalizumab in asthma and may offer potential ways to optimize asthma therapy.^16^

The varied response of many patients with severe asthma to monoclonal antibodies they are eligible for is frustrating to patients and their providers and costly to the healthcare system. Thus, identifying ways to optimize the effectiveness of these expensive therapies is of paramount importance. Many of the cells, cytokines, or their receptors targeted by these therapies are under circadian regulation. This includes the high affinity IgE receptor, FcεRI, which is downregulated by the anti-IgE, omalizumab; eosinophils, which are the primary immune cell targeted by the four anti-IL5/5R agents, and mast cell activation, which is central to immediate hypersensitivity.^17–19^ As a first step in bringing chronobiology to the field of respiratory biologics, we sought to evaluate if targets and pathways affected by respiratory biologics or related to asthma pathogenesis demonstrate significant diurnal rhythmicity. Knowing when targeted cytokines and their receptors reach peak levels may offer insights into potential time of day effects of asthma biologics.

## METHODS

### Transcriptomic datasets

We sought for relevant publicly available transcriptomic datasets in healthy controls or patients with asthma. Consistent with guidelines on genomic analyses in circadian research, we specifically looked for datasets with at most sampling every four hours with preference for every two hours sampling.^20^ Additionally, we limited to datasets that had round-the-clock (24-hour period or more) sampling to allow appropriate characterization of circadian rhythmicity.^2,20^ We found four datasets of potential relevance to this study: GSE113883, a bulk RNA sequencing study with every two hours sampling for a 28-hour period;^21^ GSE223613, a study in which healthy controls were sampled every three hours for 24 hours,^22^ and GSE196728, another bulk transcriptomics study in which participants were evaluated every four hours for 24 hours. We used the samples designated as ‘SL-Sea-Level’ which were at about 200 meters above sea level.^23^ Finally, we also identified a sleep study (GSE39445) in which participants either had sufficient sleep (control) or sleep restriction (the intervention) with samples collected every three hours for 27 hours.^24^ Transcriptomics data from GSE39445 were generated using a microarray. Thus, we selected the samples from the control condition as a cross-platform validation of our findings.

### Asthma treatment-related genes

We focused on evaluation of rhythmicity in 46 preselected genes of interest. This selection was based on: 1) their related protein being a direct or indirect target of one of the currently approved biologics (including *FCER1A, FCER1G, FCER2, IL4, IL13, IL4R, IL13RA1, IL5, IL5RA, TSLP, CRLF2 (TSLPR)*); 2.) a target currently in the asthma biologic pipeline or previously shown to be a potential target (*IL1RL1, IL25, IL33, KIT, OX40L (TNFSF4)*); 3.) an eosinophilic inflammation-associated gene or receptor (*CLC (Galectin-10), SIGLEC8*); 4.) genes potentially related to airway remodeling (*MMP9, CSF2RB, TGFB1, TIMP1*); 5) genes involved in other type-2 or effector cell biology (*GATA3, STAT6, ALOX15*); or 6) Genes related to neutrophilic inflammation and other T-helper 1 (Th-1), Th2, or Th17 inflammatory cytokines (*CXCL8, CXCL10, IL-6, IL18, IL1ß*) (**Table 1**).^25,26^

**Table 1:** The 16 clock genes and 46 preselected asthma-treatment related genes of Interest.

| ASTHMA-TREATMENT-RELATED GENES (N=46) |  |  |  |  |  |
|---|---|---|---|---|---|
| *ALOX15* | *CCL11* | *CCL17* | *CCL26* | *CCR3* | *CLC* |
| *CRLF2* (*TSLPR*) | *CSF2* | *CSF2B* | *CXCL1* | *CXCL8* | *CXCL10* |
| *FCER1A* | *FCER1G* | *FCER2* | *GATA3* | *IFNG* | *IL1B* |
| *IL1RL1 (ST2)* | *IL3* | *IL4* | *IL4R* | *IL5* | *IL5RA* |
| *IL6* | *IL10* | *IL13* | *IL13RA1* | *IL17A* | *IL17C* |
| *IL17F* | *IL18* | *IL25* | *IL33* | *KIT* | *MMP9* |
| *MS4A2* | *PRG2* | *SIGLEC8* | *STAT6* | *TGFB1* | *TIMP1* |
| *TNF* | *TNFSF4* (*OX40L*) | *TPSAB1* | *TSLP* |  |  |
| CLOCK GENES (N=16) |  |  |  |  |  |
| *ARNTL* (*BMAL1*) | *CLOCK* | *CRY1* | *CRY2* | *DBP* | *HLF* |
| *NPAS2* | *NR1D1* (*REV-ERBα*) | *NR1D2* (*REV-ERBβ*) | *PER1* | *PER2* | *PER3* |
| *RORA* | *RORB* | *RORC* | *TEF* |  |  |

### Canonical clock genes as positive controls

As positive controls, we evaluated rhythmicity in 16 circadian-rhythm related genes which included the core clock genes, *BMAL1*, *CLOCK*, and the repressors, *REV-ERB*α and *REV-ERB*β (**Table 1**).^12,14,27^ These genes are known to oscillate over the course of the 24-hour day-night cycle, and we *a priori* used them to determine if a dataset showed significant rhythmicity in at least 70% of these clock genes and should be included in further analyses looking at rhythmicity in the asthma genes.

### Data Processing and Quality Control (QC)

We downloaded each gene expression dataset from the NCBI Gene Expression Omnibus (GEO) or the relevant online repository as indicated by the authors in the primary research that generated the dataset. Thereafter, we used the metadata, which describes the study, to evaluate study records, protocol, sample description, data structure, sampling conditions, and assay platform. We identified sample IDs and sampling time using clock time to allow harmonization across datasets and identified the quality control pipeline used for each dataset. Given the uniform availability of processed post-quality control (QC) files across studies, we relied on the QC performed by the original investigators and used the study-provided processed expression data. However, we subsequently performed additional dataset- and analyses-specific QC and checks to verify subject and sampling-time information, target-gene availability, and suitability of the expression data for rhythmicity analyses (**Table 2**).

**Table 2:**
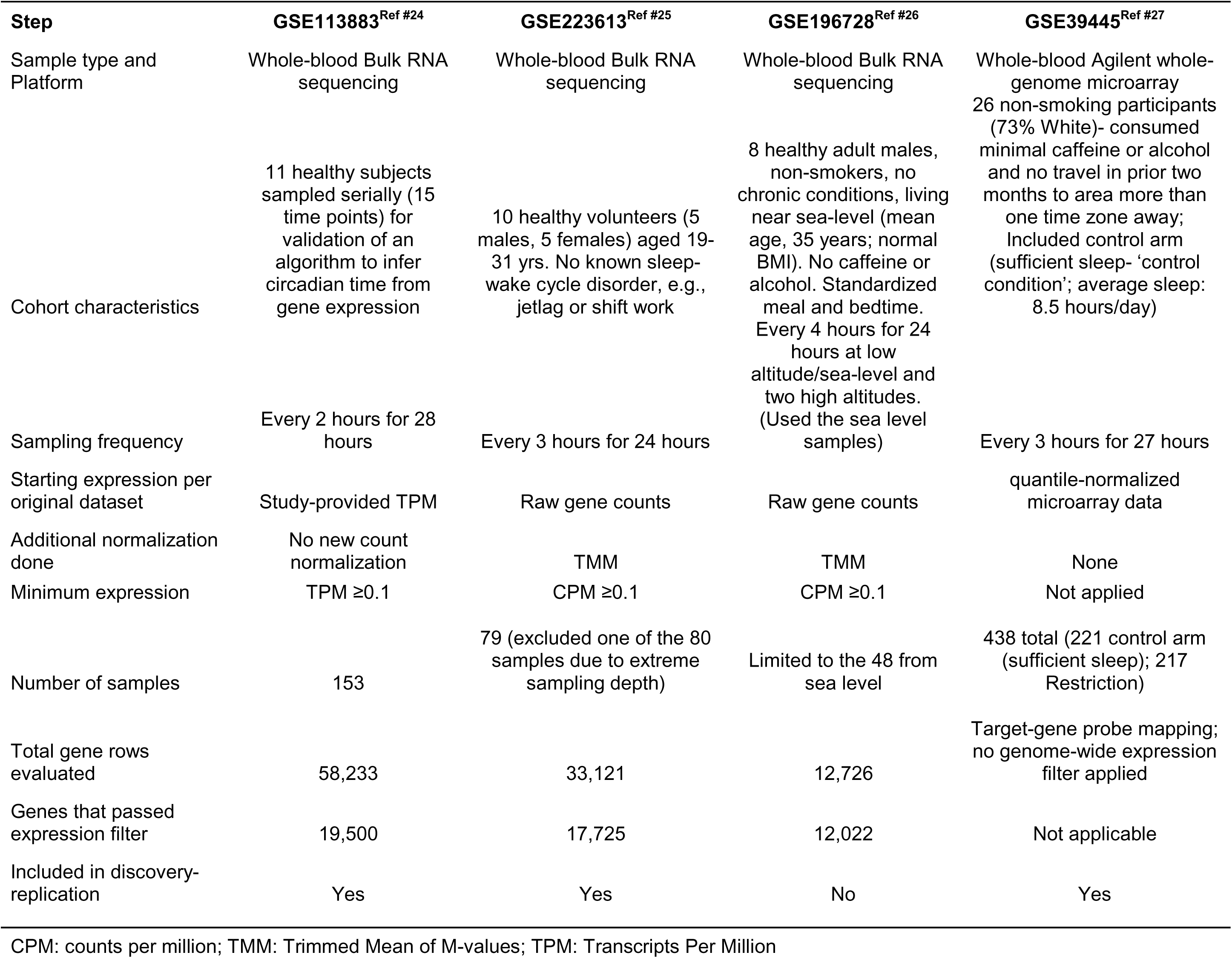
Publicly available transcriptomics datasets used in the study.

We sought to remove low-expression genes and determined if the genes of interest were ‘evaluable’ after gene expression filtering. For RNA-sequencing datasets, genes were considered evaluable if their expression was ≥0.1 TPM or ≥0.1 CPM in at least 50% of samples in the dataset. For datasets with raw gene counts, CPM values were calculated following trimmed mean of M-values (TMM) normalization. Sensitivity analyses evaluated alternative expression thresholds (TPM or CPM thresholds of 0.05, 0.1, 0.5, and 1.0 expression in at least 50% of samples). Genes failing the prespecified expression criterion were excluded from rhythmicity testing. Thereafter, we evaluated if the prespecified asthma genes and clock genes were captured in the filtered dataset. For the microarray dataset, GSE39445, we used the study-provided processed expression data, which had undergone sample-level QC and quantile normalization using the Bioconductor package limma.^24^ We first used probe annotations from the platform annotation table to map the microarray probes to the prespecified asthma genes and clock genes. Thereafter, we evaluated how many probes mapped to each target gene. For genes represented by multiple microarray probes, we conducted gene-level probe summarization by averaging the probe-level expression values within each sample to obtain a single gene-level expression value. We examined sample library sizes and identified GSM6965634 (participant J, 23:00) in GSE223613 as an extreme sequencing-depth library. We excluded that sample from the primary analytic dataset thus leaving 7 of the 8 samples for participant J in the analyses (compared to other participants sampled 8 times) and 10 samples at the eight clock times except at 23:00. Finally, given that many of the target genes of interest have other names, e.g., *CXCL8* identified as *IL8*, when a gene of interest was considered absent from a dataset, we sought for its aliases before determining that the gene was missing from the dataset.

### Statistical Analyses

For discovery-replication, we rotated the dataset used as ‘discovery’ and datasets as ‘replication’ to limit a single dataset driving our overall conclusion. We plotted gene expression over the 24-hour period for visual evaluation of rhythmicity. Thereafter, we used a 24-hour mixed-effects harmonic regression (cosinor analysis) with a subject-specific random intercept to evaluate rhythmicity of gene expression. A joint likelihood-ratio test compared this model with the random-intercept-only null model. We considered a false discovery rate (FDR) of <0.05 using the Benjamini-Hochberg test as statistical proof of rhythmicity. For every gene with significant rhythmicity, we calculated: a.) the Midline Estimating Statistic of Rhythm (MESOR), the rhythm adjusted mean, b.) the amplitude; c.) the time the gene’s expression is at its peak (acrophase); and d.) the trough time (bathyphase).^28^ Bootstrap replicates (2000 participant-level iterations) preserving within-participant correlation were used to estimate confidence intervals for these cosinor parameters. Thereafter, we evaluated *phase consistency* ‘R’ for each gene between the cohorts in which its expression was significantly rhythmic.^29^ R is the mean circular resultant length indicating the agreement of the acrophase between the cohorts, with an R of 1 indicating perfect alignment of the estimated peak times at similar clock times across the independent cohorts, and 0 meaning widely dispersed phases around the 24-hour clock. We harmonized clock time to the 24-hr clock in all datasets. Cross-dataset reproducibility was assessed among genes evaluable in at least two independent datasets. A gene was considered reproducibly rhythmic if it met FDR <0.05 in at least two independent datasets.

### Sensitivity Analyses

We evaluated if the phases of rhythmic (nominal p<0.05) clock genes were consistent with previously established phase relationships between canonical circadian clock-gene pairs. We focused on six well demonstrated relationships: *BMAL1* (core activator) antiphase to *PER1*, *PER2*, and *PER3* (core repressors) or to the transcriptional repressors, *REV-ERB*α and *REV-ERB*β.^11,30–32^ And the three core repressors in phase with each other and the transcriptional repressors in phase with each other. We limited our analysis to clock genes that replicated in at least two cohorts and with phase consistency between at least two cohorts, *R*≥0.80. To evaluate if clock gene pairs were in-phase or anti-phase, we used complimentary metrics^20,33,34^: 1) Using a sampling interval threshold, we considered genes to be in-phase if their peak times were within two hours for GSE113883 in which samples were collected every two hours, or within three hours for GSE223613 and GSE39445; 2) Using zero-lag cross-correlation of the entire wave form of the gene pairs using Pearson’s correlation coefficient. Gene pairs with maximum correlation of lag of 0 were considered in-phase; 3) Using statistical comparisons of the circular difference in the gene-pair’s acrophase and confidence intervals.

Thereafter, we fitted a two-harmonic Fourier mixed-effects model that incorporated both the fundamental 24-hour rhythm and the second 12-hour harmonic sine and cosine terms, to evaluate the robustness of our findings to alternative waveform specifications.^20^ Thereafter, to ensure our conclusions are not driven by independent participants’ influence on observed rhythms, we conducted a leave-one-participant out analyses (LOPO) and compared our results from the LOPO analyses with the original analyses. We also evaluated the impact of different gene expression filtering thresholds: TPM or CPM thresholds of 0.05, 0.1, 0.5, and 1.0 expression in at least 50% of samples. Finally, we relaxed our threshold to ≥25% of samples to evaluate if we would capture most of the filtered genes. All analyses were conducted in R version 4.5.1 and related packages (edgeR, GEOquery, dplyr, lme4, lmerTest, ggplot2, grid). ChatGPT (OpenAI) was used to assist with debugging R code. All AI-assisted code was reviewed, verified, and executed by the authors, who independently evaluated the codes and take full responsibility for the accuracy of the analyses.

## RESULTS

### Assessing rhythmicity in the canonical clock genes (positive controls)

At least 14 of the 16 (87.5%) clock genes were evaluable in all the four identified datasets. For GSE196728, though some of the 14 evaluable clock genes peaked at similar times to the other cohorts (*TEF*: 5.5h vs. 5.4 in GSE223613 and 4.1 in GSE113883; *PER1*: 6.8h vs. 6.7 vs. 6.9; *PER3*: 4.4 vs. 5.8 vs. 4.3; *REV-ERB*α: 3.6 vs. 2.0 vs. 0.9; and *CLOCK*: 23.6 vs. 23.8 vs. 3.3 respectively), only two (*PER3* and *RORA*) passed the nominal P-threshold and none passed the FDR-threshold. Thus, GSE196728 was excluded from further analyses, and we proceeded with three of the four datasets (GSE113883, GSE223613, and GSE39445) as the final discovery-replication cohorts (**Table 2**).

Fifteen of the 16 prespecified canonical clock genes were evaluable in all three cohorts (**Table 3**). All fifteen genes exhibited significant 24-hour rhythmicity in at least one dataset following multiple testing correction, 12 (80%) in at least two independent datasets, and 7 (47%) across all three datasets (**Figure 1**, **Table 3**). Phases for all replicated clock genes had high consistency between cohorts (R≥0.90) except for *DBP* (R=0.43) and *BMAL1* (*ARNTL*) (R=0.15; **Figure 1** and **Figure S1**). All replicated clock genes exhibited early morning-predominant acrophases, with a circular mean of 3.73 hours, except *BMAL1* for which the peak time in GSE113883 was 2.29h, but 15.44h in GSE223613 (**Figure 2**). The peak phase time was 14.86h in GSE39445 but did not cross the FDR threshold (p-value: 0.04; FDR: 0.08).

***Table 3:** Rhythmicity and Peak Phase of the Canonical Clock Genes.

|  | GSE113883 |  |  |  | GSE223613 |  |  |  | GSE39445<sup>#</sup> |  |  |  |  |
|---|---|---|---|---|---|---|---|---|---|---|---|---|---|
| Gene symbol | Amplit-ude | Acro-phase | Rhythmicity P-value | P-value FDR-corrected | Amplit-ude | Acro-phase | Rhythmicity P-value | P-value FDR-corrected | Amplitu-de | Acro-phase | Rhythm-icity P-value | P-value FDR-corrected | Rhythmic in # datasets |
| *HLF* | 0.067 | 3.32 | 0.015 | 0.019 | 0.202 | 4.44 | 5.87E-04 | 6.78E-04 | 0.195 | 2.53 | 6.79E-07 | 1.81E-06 | 3 |
| *REV-ERBα (NR1D1)* | 0.140 | 0.91 | 5.27E-04 | 8.78E-04 | 0.289 | 1.96 | 2.26E-13 | 1.13E-12 | 0.201 | 2.41 | 6.66E-26 | 1.07E-24 | 3 |
| *REV-ERBβ (NR1D2)* | 0.510 | 3.22 | 3.14E-15 | 2.35E-14 | 0.237 | 2.55 | 5.94E-23 | 8.91E-22 | 0.243 | 3.66 | 2.99E-25 | 2.39E-24 | 3 |
| *PER1* | 0.310 | 6.89 | 2.75E-09 | 1.03E-08 | 0.315 | 6.72 | 5.46E-09 | 1.02E-08 | 0.220 | 6.9 | 9.74E-18 | 5.19E-17 | 3 |
| *PER2* | 0.160 | 4.87 | 9.27E-09 | 2.32E-08 | 0.156 | 7.81 | 3.88E-14 | 2.91E-13 | 0.117 | 5.84 | 1.09E-08 | 3.47E-08 | 3 |
| *PER3* | 0.225 | 4.34 | 7.58E-11 | 3.79E-10 | 0.212 | 5.76 | 9.30E-10 | 2.33E-09 | 0.258 | 6.1 | 2.50E-17 | 1.00E-16 | 3 |
| *RORA* | 0.338 | 3.51 | 4.75E-09 | 1.43E-08 | 0.107 | 23.57 | 1.05E-06 | 1.43E-06 | 0.101 | 1.06 | 1.14E-03 | 2.28E-03 | 3 |
| *ARNTL (BMAL1)* | 0.135 | 2.29 | 9.34E-04 | 0.013 | 0.090 | 15.44 | 3.50E-09 | 7.50E-09 | 0.057 | 14.86 | 0.04 | 0.08 | 2 |
| *CLOCK* | 0.156 | 3.27 | 5.75E-05 | 1.08E-04 | 0.079 | 23.78 | 1.52E-07 | 2.28E-07 | 0.009 | 12.67 | 0.94 | 0.97 | 2 |
| *DBP* | 0.079 | 6.46 | 0.15 | 0.15 | 0.162 | 2.81 | 3.42E-11 | 1.03E-10 | 0.180 | 11.46 | 8.29E-06 | 1.90E-05 | 2 |
| *NPAS2* | 0.136 | 0.19 | 2.24E-03 | 3.35E-03 | 0.297 | 1.25 | 4.88E-08 | 8.14E-08 | 0.086 | 0.64 | 0.16 | 0.23 | 2 |
| *TEF* | 0.238 | 4.14 | 1.13E-16 | 1.69E-15 | 0.214 | 5.4 | 6.66E-13 | 2.50E-12 | 0.008 | 1.11 | 0.97 | 0.97 | 2 |
| *CRY1* | 0.229 | 3.54 | 3.48E-05 | 7.46E-05 | 0.016 | 0.12 | 0.71 | 0.71 | 0.043 | 5.14 | 0.27 | 0.36 | 1 |
| *CRY2* | 0.056 | 5.55 | 0.05 | 0.06 | 0.052 | 5.84 | 5.54E-05 | 6.93E-05 | 0.013 | 3.88 | 0.55 | 0.68 | 1 |
| *RORC* | 0.055 | 23.46 | 0.11 | 0.12 | 0.111 | 23.7 | 1.27E-03 | 1.36E-03 | 0.015 | 0.47 | 0.67 | 0.77 | 1 |
| *RORB* | Not evaluable |  |  |  | Not evaluable |  |  |  | 0.331 | 16.16 | 0.06 | 0.09 | 0 |

\*Acrophase (peak) in 24-hour clock; Not evaluable= filtered out
<sup>#</sup>Data from the control condition study arm

**Figure 1:**
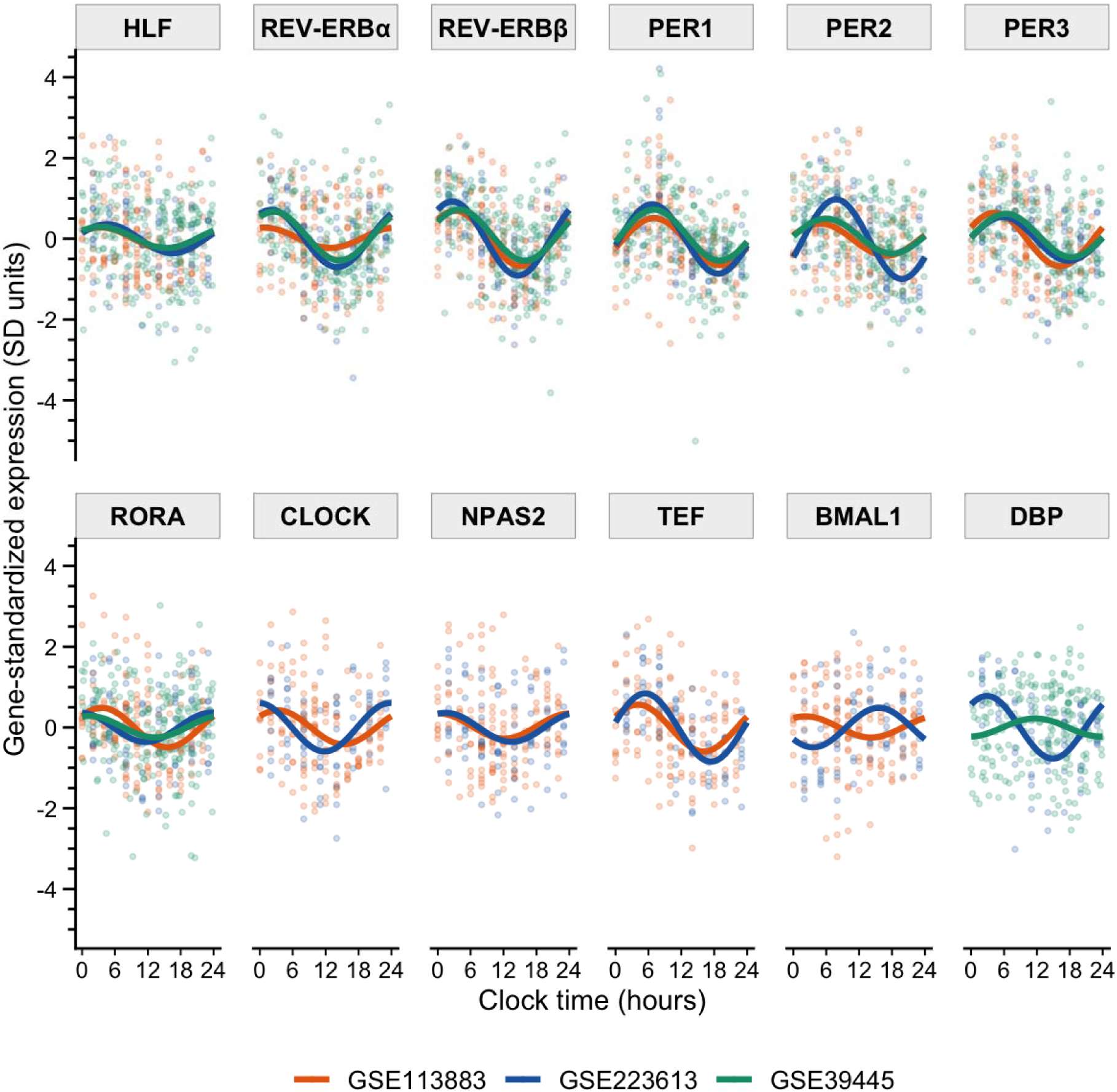
Rhythm of the 12 canonical clock genes (positive controls) showing reproducible rhythmicity (FDR<0.05) in at least 2 datasets

**Figure 2:**
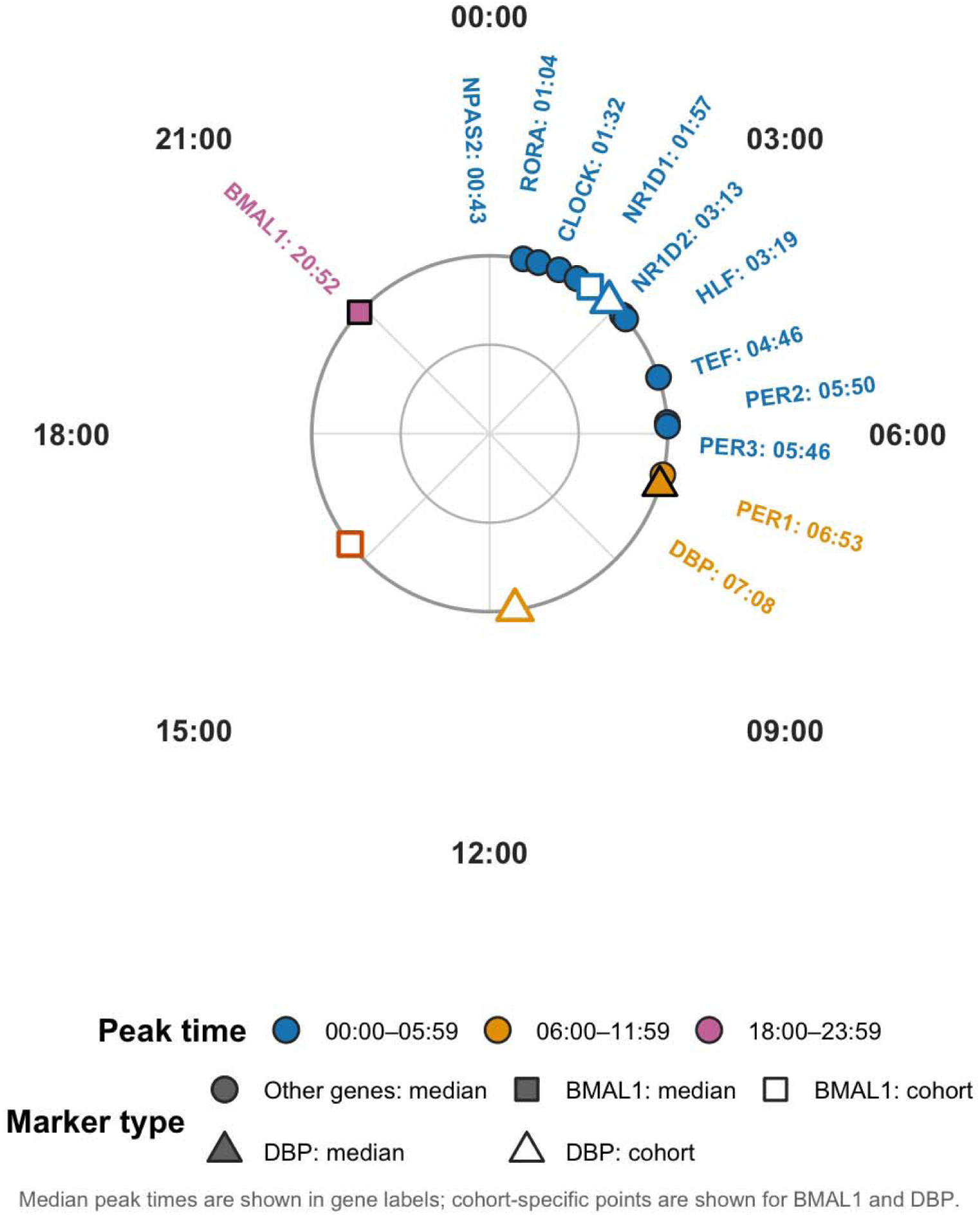
The mean peak times of the twelve replicated canonical clock genes. *Individual peak shown for genes with phase consistency, R, of <0.80*.

### Assessing rhythmicity in the prespecified asthma-treatment related genes

All 46 asthma genes were evaluable in GSE39445; 34 (74%) genes were evaluable in at least two of the three datasets, and 32 (70%) were evaluable in all three datasets (**Table 4**). Of the 34 genes evaluable in at least two of the datasets, 23 (68%) genes exhibited statistically significant and reproducible 24-hour rhythmicity (**Table 4**, **Figure 3**). Twenty-one (91%) of these genes showed high phase consistency, with R ≥80% (**Figure S1, S2**). *IL5RA* (R=0.77), *CXCL8* (R=0.67), *IL18* (R=0.53), and *ILRL1*(R=0.47), all had R of <0.80. For *IL5RA*, two of the cohorts had high acrophase consistency (GSE113883: 6.11h vs. 6.57 in GSE223613). However, GSE39445 had a phase time of 12.07 (**Table 4**, **Fig S3**). *CXCL8* was significantly rhythmic in all three datasets, but while phase was at 9.17h and 7.92h in GSE223613 and GSE39445, it was 2.82h in GSE113883. The sixteen genes that were rhythmic across all datasets were mostly phase-consistent (**Figure S2, A-C** and **Table S1**) with a median phase difference of 2.52 h (interquartile range, 1.50-4.34h) and all but one gene (*CXCL8*, shift of 6.35h) having a maximum phase difference of less than 6 hours.

***Table 4:** Rhythmicity and Peak Phase of the 46 Pre-selected Asthma-related Genes.

|  | GSE113883 |  |  | GSE223613 |  |  | GSE39445 |  |  | Evaluable in # datasets | Rhythmic in # datasets |
|---|---|---|---|---|---|---|---|---|---|---|---|
| Gene symbol | Acrophase | P-value | P-value FDR-corrected | Acrophase | P-value | P-value FDR-corrected | Acrophase | P-value | P-value FDR-corrected |  |  |
| *CLC* | 1.65 | 1.79E-06 | 7.40E-06 | 3.83 | 1.24E-03 | 2.72E-03 | 3.63 | 1.20E-07 | 6.13E-07 | 3 | 3 |
| *CXCL10* | 2.77 | 2.89E-05 | 7.96E-05 | 8.73 | 2.05E-04 | 4.83E-04 | 6.33 | 1.48E-05 | 4.87E-05 | 3 | 3 |
| *CXCL8* | 2.82 | 5.83E-07 | 3.20E-06 | 9.17 | 1.65E-03 | 3.20E-03 | 7.82 | 3.67E-04 | 8.88E-04 | 3 | 3 |
| *FCER1A* | 0.34 | 3.53E-06 | 1.06E-05 | 23.89 | 1.46E-09 | 9.64E-09 | 1.33 | 7.19E-05 | 1.94E-04 | 3 | 3 |
| *FCER1G* | 14.92 | 4.73E-07 | 3.12E-06 | 14.61 | 1.51E-03 | 3.11E-03 | 13.01 | 2.67E-10 | 4.10E-09 | 3 | 3 |
| *FCER2* | 19.82 | 2.75E-06 | 9.06E-06 | 0.34 | 2.37E-12 | 3.92E-11 | 0.65 | 1.00E-05 | 3.84E-05 | 3 | 3 |
| *GATA3* | 0.43 | 7.034E-03 | 1.055E-02 | 22.91 | 7.09E-07 | 2.60E-06 | 23.47 | 1.83E-05 | 5.62E-05 | 3 | 3 |
| *IL13RA1* | 8.62 | 6.90E-08 | 7.59E-07 | 11.69 | 1.73E-13 | 5.69E-12 | 12.79 | 4.08E-23 | 1.88E-21 | 3 | 3 |
| *IL1B* | 17.82 | 1.06E-06 | 5.00E-06 | 15.09 | 6.69E-10 | 6.00E-09 | 17.43 | 2.20E-17 | 5.07E-16 | 3 | 3 |
| *IL5RA* | 6.11 | 4.43E-09 | 7.31E-08 | 6.57 | 3.57E-03 | 6.55E-03 | 12.07 | 3.35E-04 | 8.56E-04 | 3 | 3 |
| *KIT* | 8.58 | 2.15E-06 | 7.88E-06 | 12.04 | 4.30E-03 | 7.46E-03 | 12.39 | 4.15E-08 | 2.39E-07 | 3 | 3 |
| *MMP9* | 13.26 | 4.92E-05 | 1.250E-04 | 13.29 | 1.85E-05 | 5.55E-05 | 14.03 | 1.60E-06 | 6.68E-06 | 3 | 3 |
| *SIGLEC8* | 4.06 | 4.672E-03 | 8.114E-03 | 3.32 | 6.63E-03 | 0.011 | 1.76 | 3.86E-04 | 8.89E-04 | 3 | 3 |
| *STAT6* | 13.64 | 5.737E-03 | 9.467E-03 | 13.45 | 1.92E-09 | 1.06E-08 | 15.19 | 2.78E-03 | 5.82E-03 | 3 | 3 |
| *TGFB1* | 13.25 | 1.04E-07 | 8.54E-07 | 12.68 | 1.68E-07 | 6.95E-07 | 13.35 | 1.71E-09 | 1.20E-08 | 3 | 3 |
| *TIMP1* | 14.07 | 8.08E-11 | 2.67E-09 | 14.89 | 6.32E-05 | 1.74E-04 | 14.13 | 1.12E-09 | 1.03E-08 | 3 | 3 |
| *ALOX15* | 4.03 | 3.099E-04 | 6.818E-04 | 3.91 | 1.52E-04 | 3.85E-04 | 8.37 | 0.144 | 0.221 | 3 | 2 |
| *CSF2RB* | 21.69 | 0.774 | 0.774 | 14.42 | 7.38E-09 | 3.48E-08 | 15.20 | 1.82E-09 | 1.20E-08 | 3 | 2 |
| *CXCL1* | 4.18 | 0.772 | 0.774 | 13.21 | 7.27E-10 | 6.00E-09 | 14.98 | 5.83E-07 | 2.68E-06 | 3 | 2 |
| *IL18* | 4.82 | 6.215E-03 | 9.766E-03 | 18.37 | 0.421 | 0.535 | 12.52 | 0.013 | 0.024 | 3 | 2 |
| *IL1RL1 (ST2)* | 6.67 | 3.856E-04 | 7.953E-04 | 10.77 | 0.045 | 0.065 | 14.97 | 0.014 | 0.024 | 3 | 2 |
| *IL4R* | 0.21 | 2.245E-03 | 4.359E-03 | 1.61 | 0.042 | 0.064 | 22.46 | 3.99E-05 | 1.15E-04 | 3 | 2 |
| *TNFSF4 (OX40L)* | 5.77 | 0.095 | 0.121 | 14.28 | 1.73E-05 | 5.55E-05 | 14.33 | 5.70E-10 | 6.56E-09 | 3 | 2 |
| *CCR3* | 12.12 | 0.113 | 0.138 | 11.69 | 0.084 | 0.115 | 13.91 | 1.26E-05 | 4.47E-05 | 3 | 1 |
| *IFNG* | 2.39 | 3.257E-03 | 5.971E-03 | 10.13 | 0.647 | 0.762 | 8.10 | 0.268 | 0.373 | 3 | 1 |
| *IL6* | 2.33 | 0.022 | 0.030 | 4.93 | 0.263 | 0.348 | 8.44 | 0.590 | 0.626 | 3 | 1 |
| *TNF* | 22.21 | 7.459E-03 | 0.011 | 22.37 | 0.033 | 0.052 | 21.09 | 0.224 | 0.323 | 3 | 1 |
| *TPSAB1* | 13.97 | 1.293E-04 | 3.048E-04 | 16.98 | 0.932 | 0.982 | 14.14 | 0.108 | 0.177 | 3 | 1 |
| *IL10* | 23.27 | 0.211 | 0.240 | 23.03 | 0.997 | 0.997 | 14.49 | 0.382 | 0.475 | 3 | 0 |
| *IL4* | 6.70 | 0.738 | 0.774 | 4.73 | 0.952 | 0.982 | 19.08 | 0.434 | 0.525 | 3 | 0 |
| *MS4A2 (FCER1B)* | 5.23 | 0.522 | 0.574 | 9.13 | 0.522 | 0.639 | 15.12 | 0.286 | 0.386 | 3 | 0 |
| *PRG2* | 6.82 | 0.074 | 0.097 | 6.02 | 0.904 | 0.982 | 13.45 | 0.539 | 0.604 | 3 | 0 |
| *CRLF2 (TSLPR)* | Not evaluable | Not evaluable | Not evaluable | 15.37 | 0.670 | 0.762 | 13.26 | 0.141 | 0.221 | 2 | 0 |
| *IL17C* | 5.92 | 0.144 | 0.170 | Not evaluable | Not evaluable | Not evaluable | 15.16 | 0.322 | 0.423 | 2 | 0 |
| *CCL26* | Not evaluable | Not evaluable | Not evaluable | Not evaluable | Not evaluable | Not evaluable | 18.00 | 1.10E-03 | 2.41E-03 | 1 | 1 |
| *IL17A* | Not evaluable | Not evaluable | Not evaluable | Not evaluable | Not evaluable | Not evaluable | 14.18 | 4.50E-03 | 9.01E-03 | 1 | 1 |
| *IL17F* | Not evaluable | Not evaluable | Not evaluable | Not evaluable | Not evaluable | Not evaluable | 13.50 | 6.65E-03 | 0.013 | 1 | 1 |
| *IL33* | Not evaluable | Not evaluable | Not evaluable | Not evaluable | Not evaluable | Not evaluable | 10.34 | 0.024 | 0.042 | 1 | 1 |
| *CCL11* | Not evaluable | Not evaluable | Not evaluable | Not evaluable | Not evaluable | Not evaluable | 15.12 | 0.599 | 0.626 | 1 | 0 |
| *CCL17* | Not evaluable | Not evaluable | Not evaluable | Not evaluable | Not evaluable | Not evaluable | 15.20 | 0.684 | 0.689 | 1 | 0 |
| *CSF2* | Not evaluable | Not evaluable | Not evaluable | Not evaluable | Not evaluable | Not evaluable | 23.75 | 0.335 | 0.428 | 1 | 0 |
| *IL13* | Not evaluable | Not evaluable | Not evaluable | Not evaluable | Not evaluable | Not evaluable | 1.38 | 0.689 | 0.689 | 1 | 0 |
| *IL25* | Not evaluable | Not evaluable | Not evaluable | Not evaluable | Not evaluable | Not evaluable | 21.81 | 0.564 | 0.618 | 1 | 0 |
| *IL3* | Not evaluable | Not evaluable | Not evaluable | Not evaluable | Not evaluable | Not evaluable | 23.04 | 0.490 | 0.563 | 1 | 0 |
| *IL5* | Not evaluable | Not evaluable | Not evaluable | Not evaluable | Not evaluable | Not evaluable | 6.53 | 0.472 | 0.557 | 1 | 0 |
| *TSLP* | Not evaluable | Not evaluable | Not evaluable | Not evaluable | Not evaluable | Not evaluable | 21.54 | 0.211 | 0.313 | 1 | 0 |

*\*Acrophase (peak) in 24-hour clock*
<sup>#</sup>*P-values are evaluating if significant rhythmicity in rhythm (comparing null model to model with sine-cosine values); FDR- Benjamini-Hochberg False Discovery Rate*

**Figure 3:**
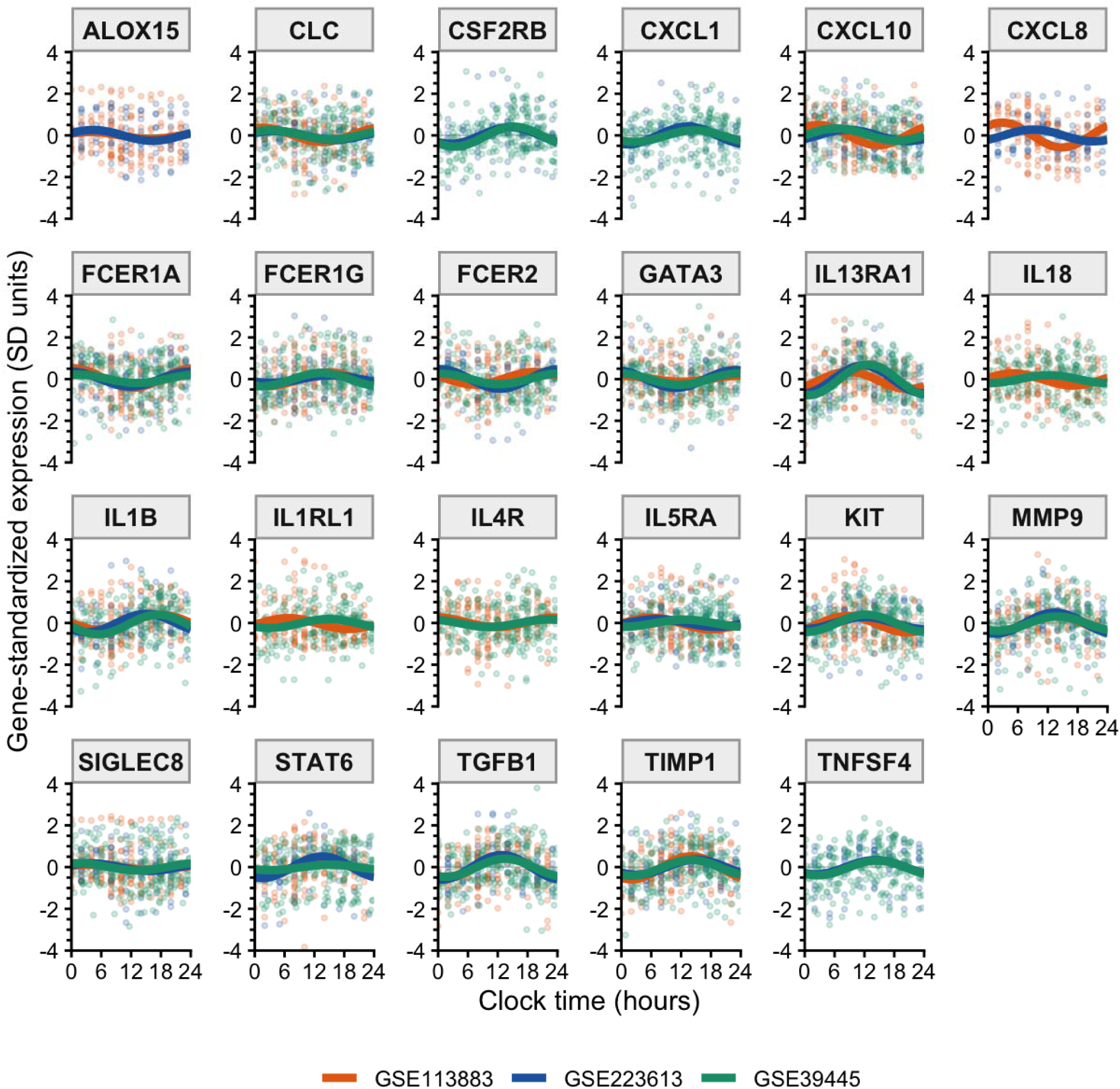
Rhythm of the 23-asthma therapy-related genes showing reproducible rhythmicity (FDR<0.05) in at least 2 datasets.

Using the circular mean peak time of the 23 reproducibly rhythmic asthma genes across cohorts in which the gene was statistically significant, seven genes, including *FceR1A*, *FcER2, ALOX15, GATA3*, *and IL4R* peaked in the late night or early period of the morning (23.00 to 04.00h), seven peaked in the early morning (~06.00h to ~11.00h period), including *IL5RA*, *IL1RL1*, *KIT*, and *IL13RA1* (**Figure 4; Table S2**). The other nine genes peaked in the afternoon to early evening (13.00 to 18.00h). This group included airway remodeling associated genes: *TGFB1*, *MMP9*, and *TNFSF4* (*OX40L*).

**Figure 4:**
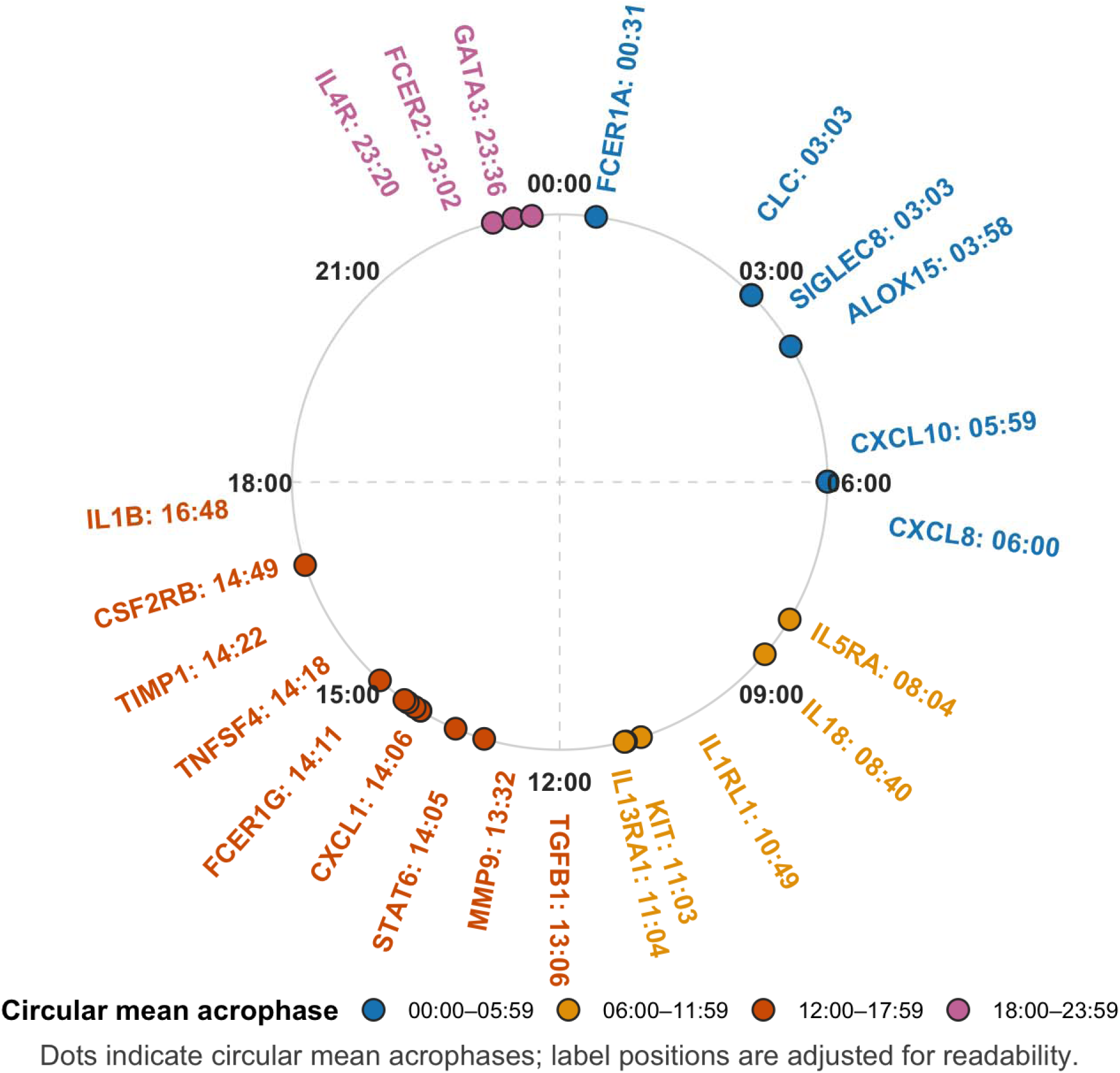
The circular mean peak time of the 23 asthma-therapy related genes with reproducible 24-hour rhythmicity across cohorts.

### Sensitivity Analyses

We evaluated if known phase relationships between clock gene pairs were preserved in 10 of the 16 clock genes that were significant at the nominal threshold and exhibited high phase consistency between at least two of the three datasets. These included *BMAL1*, which though had low cross-cohort phase consistency across the three datasets had a high phase consistency (0.997) between GSE223613 and GSE39445 (**Figure S1**). *BMAL* was antiphase to both *REV-ERB*α and *REV-ERB*β with both these transcriptional repressors in phase with each other. Across the three cohorts *PER1*, *PER2*, and *PER3* were also in phase (**Table S3**).

We compared the results of our primary 24-h harmonic cosinor model with that of the Fourier two-harmonic (12- and 24-hour) mixed-effects model. Results were similar with 22 (96%) of the 23 genes demonstrating cross-dataset replication (**Figure S4**). *IL18* lost replication and *CCR3* and *TNF,* which were significantly rhythmic in only one dataset in our primary analyses, became reproducible with two datasets showing significant rhythmicity in the two-harmonic Fourier model (**Table 4**, **Figure S4**).

In Leave-One-Participant-Out (LOPO) analyses, 19 of 20 (95%) genes that were significantly rhythmic in GSE113883 retained their significance. The largest acrophase difference was 1.07h (64 minutes). *STAT6* was not significant (p=0.08) in LOPO analyses. In GSE34995, all 21 genes replicated in the primary analyses were significantly rhythmic in LOPO analyses, with a largest acrophase difference of 0.90h (54 minutes). In GSE223613, 19 of 20 (95%) genes significantly rhythmic in the primary model retained their significance. The largest acrophase difference was 1.23h. *SIGLEC8* was the only gene that lost significance (p=0.14) in LOPO analyses of GSE223613 (**Table S4**).

When we changed the thresholds of expression filtering from TPM≥0.10 or CPM≥0.10 to ≥0.01, 0.05, 0.50, and 1.00, 18 (90%) of the 20 replicated genes from primary analyses of GSE113883 retained significant rhythmicity (FDR <0.05) and 20 (100%) of the 20 rhythmic genes from GSE223613 retained their rhythmicity. For GSE113883, all 23 replicated asthma genes remained evaluable at TPM thresholds of 0.01, 0.05, and 0.10, with 20 retaining significant rhythmicity. At more stringent thresholds of 0.50 and 1.00 TPM, 21 and 20 replicated genes remained evaluable, respectively, with 18 retaining significant rhythmicity at both thresholds. The two genes lost were excluded due to the stringent expression filters (*KIT* and *IL1RL1*), rather than loss of statistical significance among retained genes. In GSE223613, all 23 replicated asthma genes remained evaluable across CPM thresholds of 0.05-1.00, and all 20 originally rhythmic genes retained significance (**Table S5**). *RORB* and *IL33* passed the threshold filter in ≥25% of samples in the GSE223613 cohort (39% and 38% of samples respectively). However, neither showed significant rhythmicity in this cohort (p= 0.088 and 0.203 respectively, **Table S6**)

## DISCUSSION

In this study using transcriptomics datasets with round-the-clock sampling, we evaluated if genes associated with receptors or pathways targeted by asthma treatment show significant 24-h variability. Three-quarters of the canonical clock genes evaluable in the datasets showed significant rhythmicity. Clock genes all peaked mostly in the night and early morning and showed high phase consistency (R ≥0.90), except DBP (R=0.43) and *BMAL* which showed low cross-cohort consistency with GSE113883, but high consistency (R=0.997) between GSE223613 and GSE39445. As expected, in GSE223613 and GSE39445, *BMAL1* oscillated antiphase to the core repressors (*PER1, PER2, PER3*) and to the nuclear receptors, *REV-ERB*α and *REV-ERB*β. We found that a high proportion of genes associated with various receptors or pathways targeted by current or potential future therapies for asthma demonstrate significant 24-h rhythmicity. Of the 34 genes evaluable in two or more cohorts, 23 demonstrated significant rhythmicity at the Benjamini-Hochberg corrected threshold. These included signal transduction factors like *GATA3* and *STAT6*, master regulators of type 2 inflammation and IL4/IL13 signaling,^35,36^ chemokines, including the neutrophil chemoattractant, *CXCL8*, and surface receptors, including the high affinity IgE receptor, which is suppressed by the anti-IgE agent, omalizumab; *IL4R* and *IL13RA1*, blocked by dupilumab, and *IL5RA*, blocked by the anti-IL5 receptor agent, benralizumab. There was high phase consistency between most of the replicated genes and our conclusions were robust to waveform specification, varying gene expression thresholds, and in leave-one-participant out analyses.

To date, seven respiratory biologics are approved for the treatment of asthma with dozens in the drug development pipeline.^25,37,38^ While these therapies have shown high efficacy in clinical trials, up to 40% or more of individuals eligible for these biologics in everyday clinics have suboptimal response.^39–41^ This delay in asthma control is frustrating to patients and their providers and costly to the healthcare system. Finding ways to optimize these therapies is of utmost importance. There is emerging evidence that the anti-IgE, omalizumab, might demonstrate some time-of-day variation in its effect.^16^ The high affinity IgE receptor, FcεRI, which is downregulated by the anti-IgE, omalizumab, is under strict circadian regulation and disrupted and/or mutated core circadian pathways can abolish IgE-mediated anaphylaxis in mice.^17,18^ In this study, we found this high affinity receptor and it’s low affinity counterpart (*FcεRII*) to have strong rhythmicity peaking at around midnight. Given the important role the circadian rhythm plays in inflammatory processes, the endogenous circadian nature of asthma,^2^ and the imperfect response to biologics even in patients who meet eligibility, understanding the 24-h variation in expression of these and other targeted receptors or pathways can be potentially practice-changing leading to therapy optimization. *IL4R* expression peaked at almost midnight, and *IL5RA* expression peaked at about 8 AM suggesting that late nighttime dosing of dupilumab and morning dosing of benralizumab might be best. However, additional research is needed on how peak expression of these receptors impacts therapeutic benefit.

Our finding that many of the asthma-related genes evaluated showed significant daily rhythmicity are unsurprising given that over forty percent of the mammalian genome is under circadian clock control.^11^ The endogenous circadian timing system influences many cellular functions involved in asthma pathogenesis, allergic inflammation, tissue remodeling, and cytokine expression.^2,12–14,42^ Asthma is in itself a rhythmic disease showing oscillations in severity and control with late night and early morning worsening of symptoms.^3,5^ Furthermore, just like the influence of the circadian system on asthma severity, asthma control and airway inflammation can be incited or disrupted by multiple environmental factors such as light exposure, hormones, a common explanation for the higher prevalence of asthma in adult females, and exogenous glucocorticoids.^1,3,43^ These suggest asthma as a prime area for chronotherapy. In addition, ample research has shown that the lung has its own intrinsic circadian rhythm.^11,14,27^ The lung’s inflammatory response to lipopolysaccharide (LPS) and subsequent CXCL5 expression and pulmonary neutrophil recruitment, for example, depends on the time of day.^44^ Likewise, effectiveness of glucocorticoids requires an intact airway circadian clock,^44,45^ and many pathogenic processes involved in asthma, including allergic inflammation and tissue remodeling, are all under circadian influence.^17,44–47^ Harnessing these potential time-of-day effects have been used across chronic inflammatory diseases, like rheumatoid arthritis, and cancer, and may offer potential ways to optimize asthma therapy.^15,48–50^

Our results have multiple limitations. We used publicly available blood-based transcriptomics datasets and some of these were pre-processed. Though the high phase consistency between independent cohorts suggest that this is unlikely to explain our results, phases of clock genes or of the asthma-related transcriptome may differ across blood, cells, and organs.^11,51^ Furthermore, the range of rhythmic targets found suggest that, beyond surface receptors, underlying signal transduction processes themselves can be under circadian influence. Secondly, these studies included samples collected over 24-28-hour periods. However, at least two complete cycles (≥48 hours) would be preferred in establishing rhythmicity.^20^ Thirdly, the rhythmicity observed here might be related to rhythmicity in the abundance of cells expressing these genes, not a reflection of the expression of genes per cells themselves, given that these were assessed in circulating blood not at the cellular level. Nonetheless, our results show that many targets in the treatment of asthma show diurnal rhythmicity and targeting these, or their cells of origin, may exhibit possible time of day therapeutic effects. We harmonized and used local clock time. However, the chronotype of these research participants and their physiologic times may differ and aligning the data relative to each individual’s endogenous circadian time, e.g., dim light melatonin onset (DLMO) as a marker of the central circadian clock, might provide different results.^52^ Furthermore, most of the rhythmic genes were genes associated with receptors. Many of the genes associated with T2 cytokines, *IL4*, *IL5*, *IL13*, *TSLP*, did not demonstrate significant rhythmicity likely because they were not evaluable in all the cohorts and may have had low expression in circulating blood. Thus, future studies should evaluate and use the best compartments in which to measure these. Finally, these data sets were in healthy controls. Although these would reflect fundamental patterns in human biology, these pathways are likely dysregulated in individuals with asthma and thus our results might not translate directly to patients with asthma.

In conclusion, we found that gene expression of many of the therapeutic targets currently approved for asthma biologics or in the drug development pipeline exhibit significant 24-h rhythmicity. Future studies should evaluate time of day effects of targeted therapies in asthma.

## Data Availability

All datasets are available online at NCBI Gene Expression Omnibus (GEO).

## CONFLICTS OF INTEREST

AA and VJC report no relevant conflicts. FAJLS has received consulting fees from the University of Alabama at Birmingham, Morehouse School of Medicine, and Salk Institute for Biological Studies. FAJLS interests were reviewed and managed by Brigham and Women’s Hospital and Mass General Brigham in accordance with their conflict-of-interest policies. FAJLS consultancies are not related to the current work. STW receives royalties from UpToDate.

## FUNDING & ACKNOWLEDGEMENT

This work was conducted with support from the American Lung Association (ALA)/American Academy of Allergy, Asthma & Immunology (AAAAI) Allergic Respiratory Diseases Award and the UM1TR004408 award through Harvard Catalyst Consulting Service | The Harvard Clinical and Translational Science Center (National Center for Advancing Translational Sciences, National Institutes of Health) and financial contributions from Harvard University and its affiliated academic healthcare centers. The content is solely the responsibility of the authors and does not necessarily represent the official views of Harvard Catalyst, Harvard University and its affiliated academic healthcare centers, or the National Institutes of Health.

## Notes

### Author Declarations

We used relevant publicly available transcriptomic datasets that were originally located at the NCBI Gene Expression Omnibus (GEO): 1. GSE223613: https://www.ncbi.nlm.nih.gov/geo/query/acc.cgi?acc=GSE223613 2.GSE113883: https://www.ncbi.nlm.nih.gov/geo/query/acc.cgi?acc=GSE113883 3.GSE39445: https://www.ncbi.nlm.nih.gov/geo/query/acc.cgi?acc=GSE39445 4. GSE196728: https://www.ncbi.nlm.nih.gov/geo/query/acc.cgi?acc=GSE196728

### Summary of Updates

Changing license from restricted to CC-BY in compliance with NIH's Open Access Policy

