## Supplemental File for "Timing the Target: Exploring the Circadian Transcriptome of Biologic Therapies in Asthma"

**SUPPLEMENTARY FIGURES AND TABLES**

**Figure S1:** Cross-cohort consistency of 24-hour peak phase (acrophase) in genes reproducibly rhythmic in two or more evaluable cohorts. *R ranges from 0 to 1 with higher values indicating greater cross-cohort agreement of the peak phase. Dashed lines indicate the median R within each gene set.*

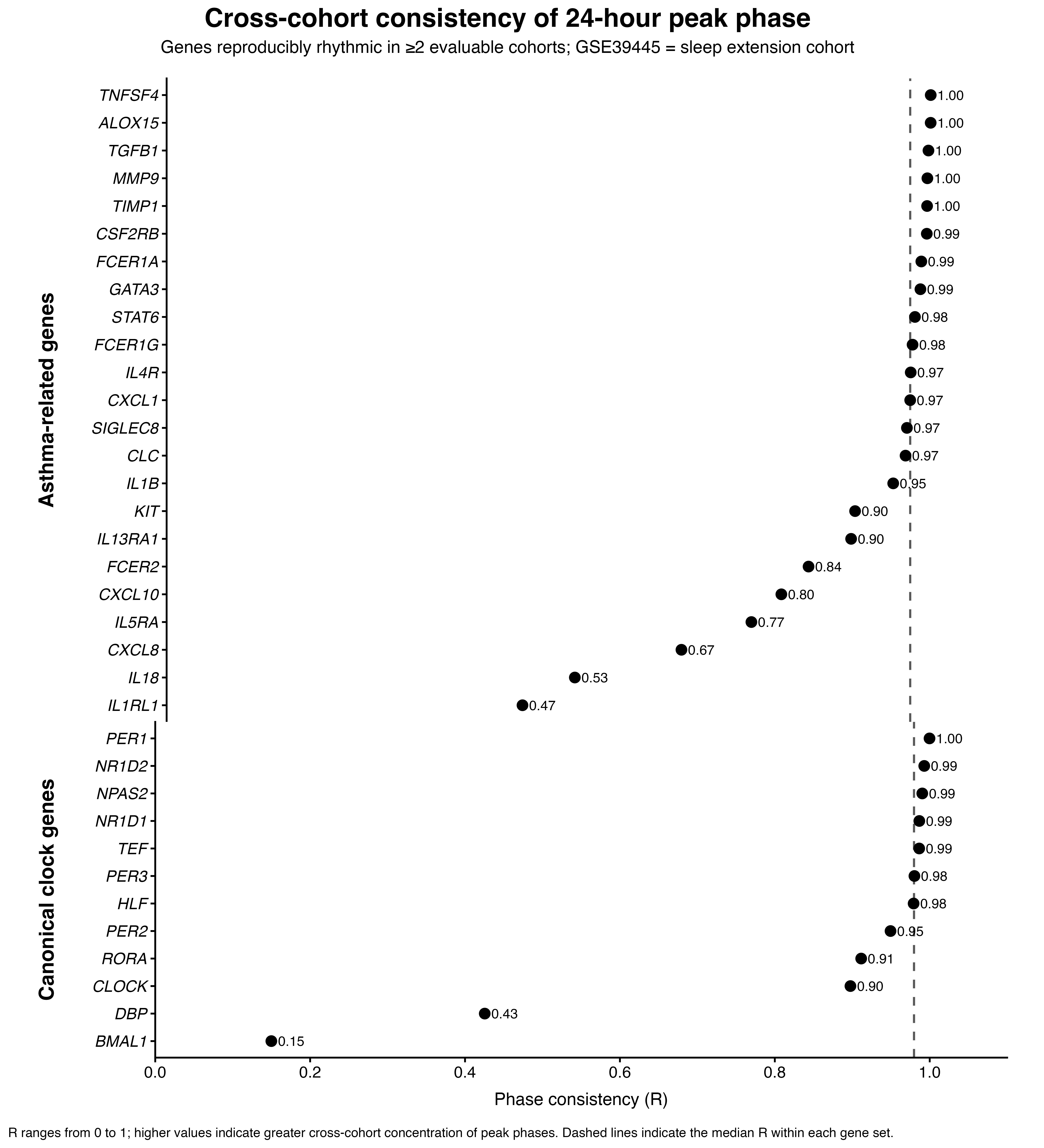

**Figure S2**: Acrophase (24-hour peak phase) of the 23 reproducibly rhythmic asthma-related genes across cohorts.

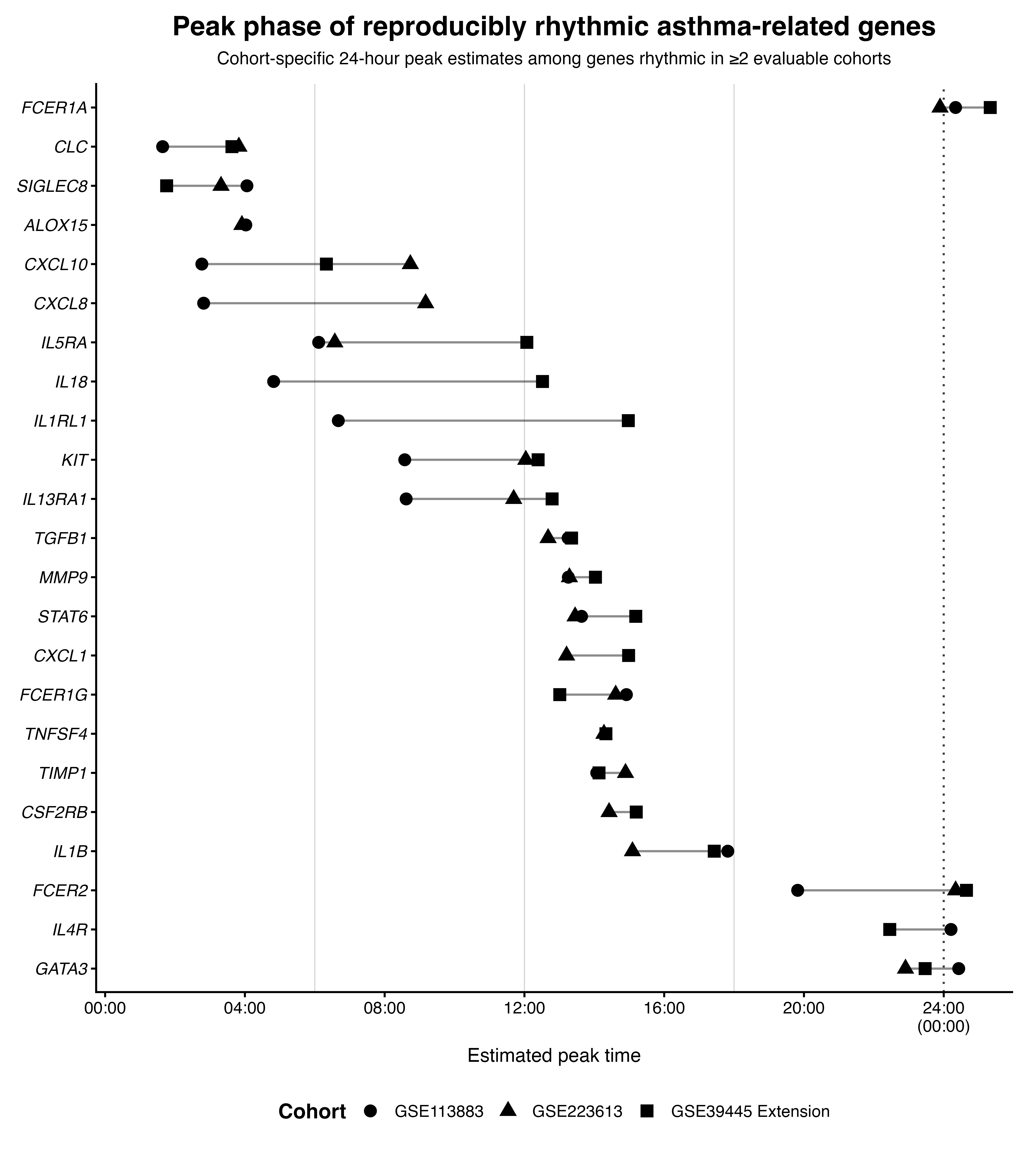

**Figure S3, A-C**: Acrophase (peak phase time) of the 23-asthma therapy-related genes with significant rhythmicity

**A.**

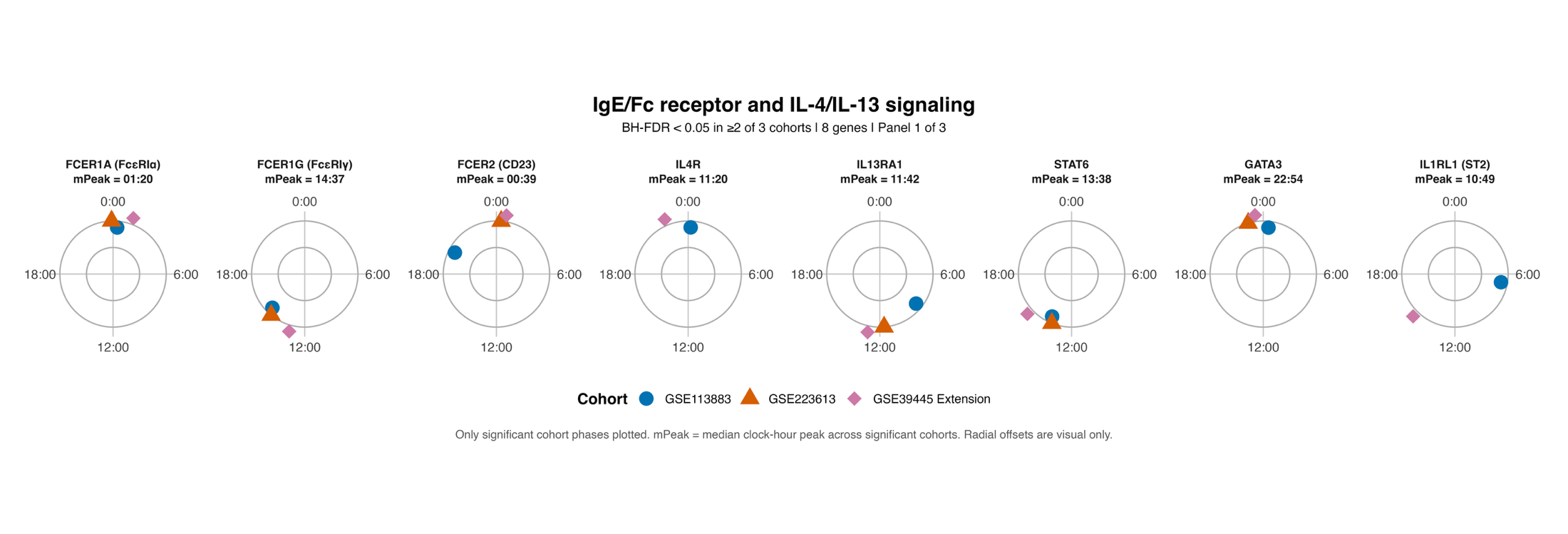

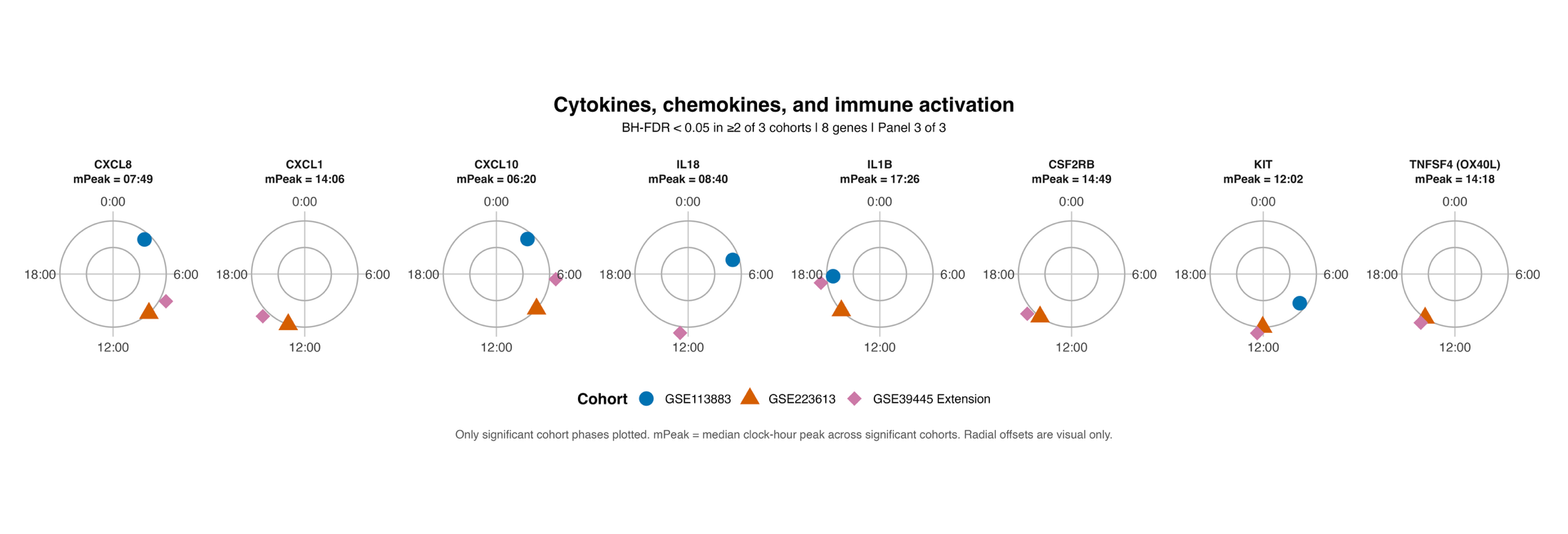

**C.**

**B.**

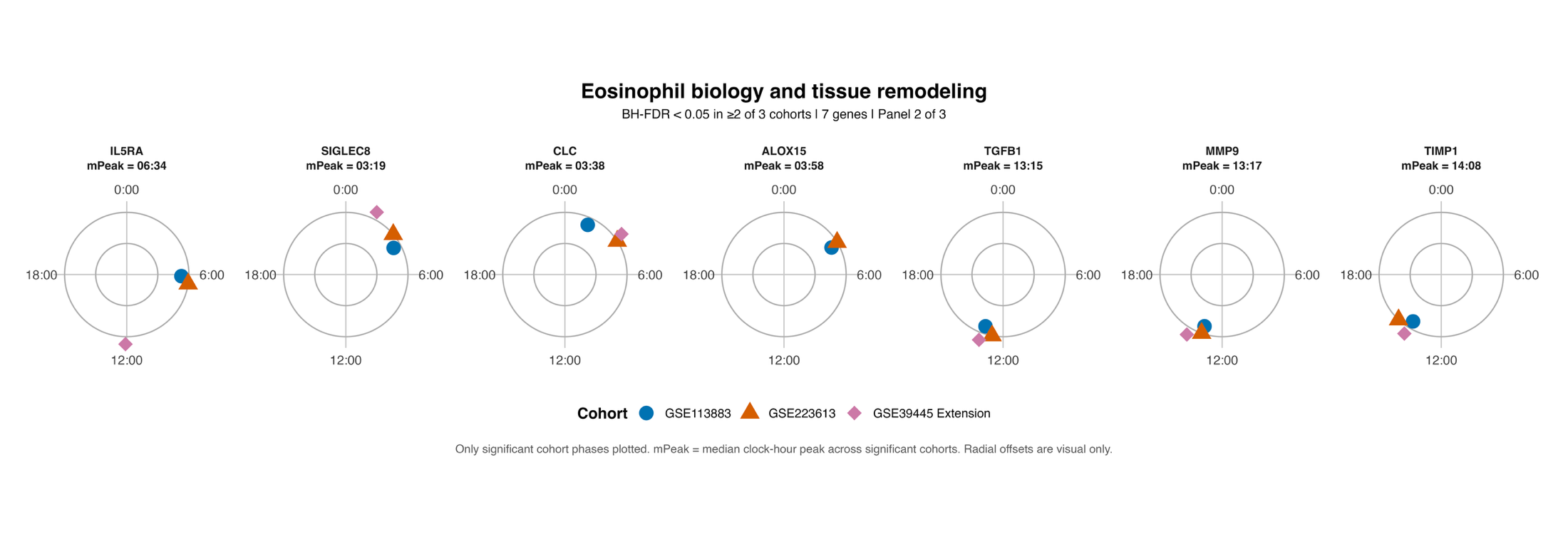

**Figure S4**: Cosinor and Fourier fitted expression for the four genes with discordant results from cosinor vs. Fourier analyses

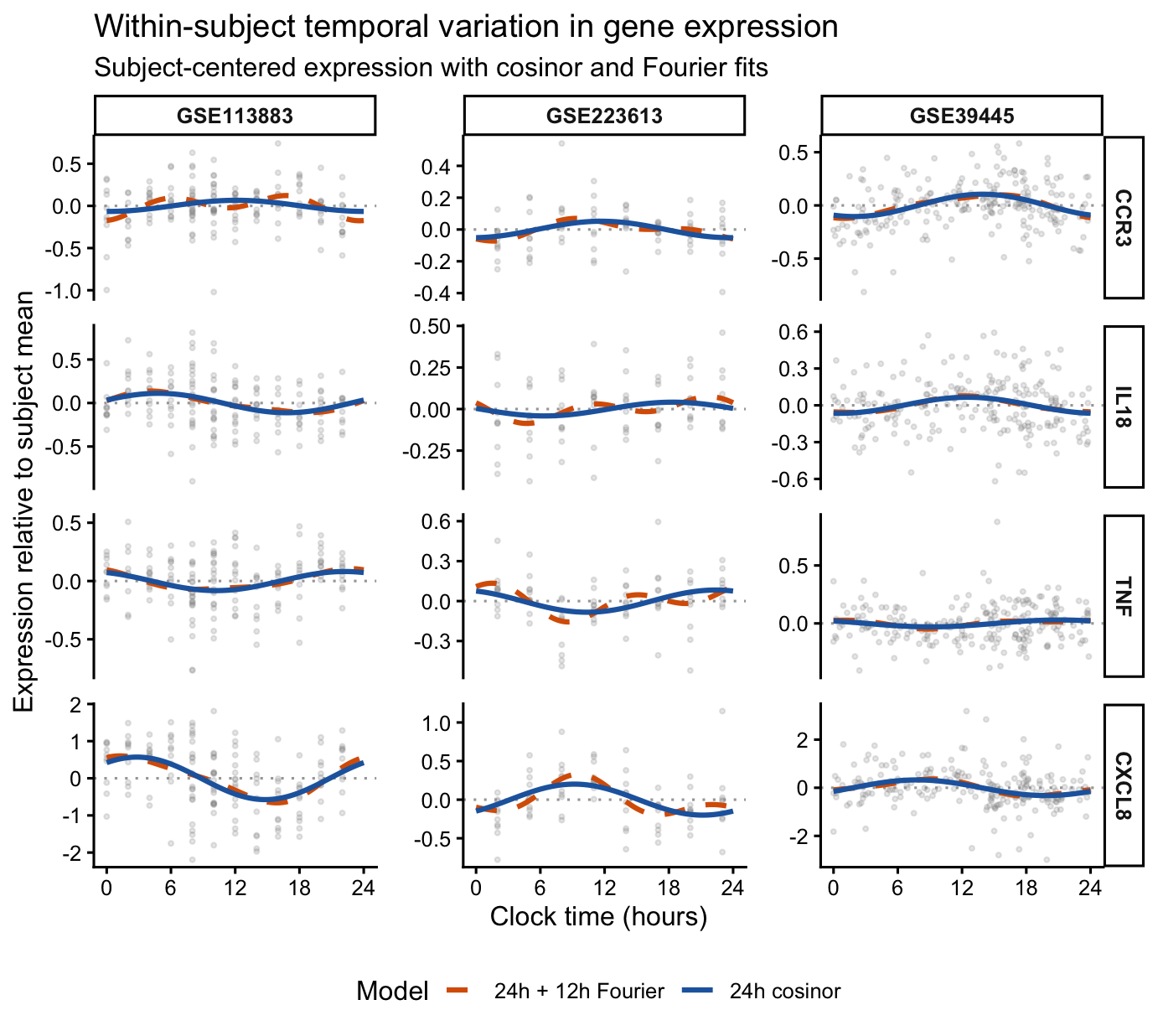

**Table S1**: Circular mean differences over 24-hr clock time in acrophase (peak time) between cohorts in the 16 asthma genes evaluable and rhythmic in all three datasets.

| **Gene Symbol** | **Acrophase (peak time)** | | | **Circular difference between the peak times** | | | **Maximum circular difference** |
| --- | --- | --- | --- | --- | --- | --- | --- |
|  | GSE113883 | GSE223613 | GSE39445 | 113883 vs. 223613 | 113883 vs. 39445 | 223613 vs. 39445 |  |
| *CLC* | 1.65 | 3.83 | 3.63 | 2.18 | 1.98 | 0.2 | 2.18 |
| *CXCL10* | 2.77 | 8.73 | 6.33 | 5.96 | 3.56 | 2.4 | 5.96 |
| *CXCL8* | 2.82 | 9.17 | 7.82 | 6.35 | 5 | 1.35 | 6.35 |
| *FCER1A* | 0.34 | 23.89 | 1.33 | 0.45 | 0.99 | 1.44 | 1.44 |
| *FCER1G* | 14.92 | 14.61 | 13.01 | 0.31 | 1.91 | 1.6 | 1.91 |
| *FCER2* | 19.82 | 0.34 | 0.65 | 4.52 | 4.83 | 0.31 | 4.83 |
| *GATA3* | 0.43 | 22.91 | 23.47 | 1.52 | 0.96 | 0.56 | 1.52 |
| *IL13RA1* | 8.62 | 11.69 | 12.79 | 3.07 | 4.17 | 1.1 | 4.17 |
| *IL1B* | 17.82 | 15.09 | 17.43 | 2.73 | 0.39 | 2.34 | 2.73 |
| *IL5RA* | 6.11 | 6.57 | 12.07 | 0.46 | 5.96 | 5.5 | 5.96 |
| *KIT* | 8.58 | 12.04 | 12.39 | 3.46 | 3.81 | 0.35 | 3.81 |
| *MMP9* | 13.26 | 13.29 | 14.03 | 0.03 | 0.77 | 0.74 | 0.77 |
| *SIGLEC8* | 4.06 | 3.32 | 1.76 | 0.74 | 2.3 | 1.56 | 2.3 |
| *STAT6* | 13.64 | 13.45 | 15.19 | 0.19 | 1.55 | 1.74 | 1.74 |
| *TGFB1* | 13.25 | 12.68 | 13.35 | 0.57 | 0.1 | 0.67 | 0.67 |
| *TIMP1* | 14.07 | 14.89 | 14.13 | 0.82 | 0.06 | 0.76 | 0.82 |

| **Table S2**: **Cosinor Parameters (across 24h) for the 23 Asthma Genes that Replicated across Cohorts** | | | | | |
| --- | --- | --- | --- | --- | --- |
| Gene symbol | Cohort | *^#^MESOR (95% CI) (cohort-specific log2-expression units) | *^#^Amplitude, 95% CI; (cohort-specific log2-expression units) | Acrophase (peak clock-hr), 95% CI | Bathyphase (trough clock-hr), 95% CI |
| ALOX15 | GSE113883 | 3.63 (2.97‚ 4.39) | 0.240 (0.097‚ 0.395) | 4.03 (1.99‚ 7.04) | 16.03 (13.99‚ 19.04) |
| ALOX15 | GSE223613 | 5.20 (4.69‚ 5.69) | 0.240 (0.085‚ 0.449) | 3.91 (2.71‚ 6.11) | 15.91 (14.71‚ 18.11) |
| ALOX15 | GSE39445 | 5.18 (4.99‚ 5.36) | 0.054 (0.026‚ 0.100) | 8.37 (5.43‚ 12.70) | 20.37 (17.43‚ 0.70) |
| CLC | GSE113883 | 8.32 (7.85‚ 8.81) | 0.346 (0.197‚ 0.525) | 1.65 (23.30‚ 3.24) | 13.65 (11.30‚ 15.24) |
| CLC | GSE223613 | 6.42 (6.00‚ 6.89) | 0.164 (0.044‚ 0.293) | 3.83 (2.82‚ 5.41) | 15.83 (14.82‚ 17.41) |
| CLC | GSE39445 | 12.79 (12.49‚ 13.09) | 0.189 (0.128‚ 0.265) | 3.63 (2.04‚ 5.58) | 15.63 (14.04‚ 17.58) |
| CSF2RB | GSE113883 | 6.60 (6.25‚ 6.95) | 0.038 (0.015‚ 0.169) | 21.69 (11.62‚ 7.51) | 9.69 (23.62‚ 19.51) |
| CSF2RB | GSE223613 | 10.05 (9.80‚ 10.29) | 0.182 (0.105‚ 0.267) | 14.42 (13.66‚ 15.54) | 2.42 (1.66‚ 3.54) |
| CSF2RB | GSE39445 | 9.26 (9.14‚ 9.38) | 0.226 (0.167‚ 0.301) | 15.20 (13.27‚ 16.76) | 3.20 (1.27‚ 4.76) |
| CXCL1 | GSE113883 | 2.98 (2.89‚ 3.09) | 0.032 (0.013‚ 0.154) | 4.18 (16.87‚ 14.88) | 16.18 (4.87‚ 2.88) |
| CXCL1 | GSE223613 | 3.21 (2.88‚ 3.56) | 0.275 (0.184‚ 0.362) | 13.21 (12.37‚ 13.93) | 1.21 (0.37‚ 1.93) |
| CXCL1 | GSE39445 | 10.24 (10.00‚ 10.45) | 0.177 (0.110‚ 0.249) | 14.98 (13.80‚ 16.19) | 2.98 (1.80‚ 4.19) |
| CXCL10 | GSE113883 | 1.21 (1.03‚ 1.39) | 0.260 (0.189‚ 0.330) | 2.77 (1.20‚ 4.15) | 14.77 (13.20‚ 16.15) |
| CXCL10 | GSE223613 | 0.50 (-0.10‚ 1.13) | 0.328 (0.217‚ 0.469) | 8.73 (6.53‚ 9.98) | 20.73 (18.53‚ 21.98) |
| CXCL10 | GSE39445 | 3.68 (3.24, 4.11) | 0.348 (0.212, 0.528) | 6.33 (3.92, 8.37) | 18.33 (15.92, 20.37) |
| CXCL8 | GSE113883 | 2.55 (2.32‚ 2.78) | 0.572 (0.450‚ 0.710) | 2.82 (1.29‚ 4.12) | 14.82 (13.29‚ 16.12) |
| CXCL8 | GSE223613 | 3.93 (3.56‚ 4.30) | 0.201 (0.086‚ 0.351) | 9.17 (6.86‚ 11.47) | 21.17 (18.86‚ 23.47) |
| CXCL8 | GSE39445 | 5.52 (4.76, 6.16) | 0.325 (0.203, 0.472) | 7.82 (5.90, 9.68) | 19.82 (17.90, 21.68) |
| FCER1A | GSE113883 | 5.21 (4.91‚ 5.53) | 0.406 (0.302‚ 0.531) | 0.34 (22.56‚ 1.76) | 12.34 (10.56‚ 13.76) |
| FCER1A | GSE223613 | 4.73 (4.46‚ 5.00) | 0.163 (0.098‚ 0.232) | 23.89 (23.31‚ 0.60) | 11.89 (11.31‚ 12.60) |
| FCER1A | GSE39445 | 10.50 (10.26‚ 10.72) | 0.158 (0.088‚ 0.245) | 1.33 (23.00‚ 3.55) | 13.33 (11.00‚ 15.55) |
| FCER1G | GSE113883 | 9.93 (9.64‚ 10.23) | 0.159 (0.091‚ 0.232) | 14.92 (13.19‚ 16.25) | 2.92 (1.19‚ 4.25) |
| FCER1G | GSE223613 | 5.85 (5.59‚ 6.17) | 0.094 (0.042‚ 0.163) | 14.61 (13.40‚ 18.24) | 2.61 (1.40‚ 6.24) |
| FCER1G | GSE39445 | 14.41 (14.16‚ 14.65) | 0.234 (0.160‚ 0.322) | 13.01 (12.11‚ 14.16) | 1.01 (0.11‚ 2.16) |
| FCER2 | GSE113883 | 5.18 (4.90‚ 5.48) | 0.180 (0.110‚ 0.260) | 19.82 (17.94‚ 21.67) | 7.82 (5.94‚ 9.67) |
| FCER2 | GSE223613 | 4.49 (4.17‚ 4.74) | 0.259 (0.194‚ 0.331) | 0.34 (0.11‚ 0.52) | 12.34 (12.11‚ 12.52) |
| FCER2 | GSE39445 | 6.35 (6.12‚ 6.58) | 0.139 (0.089‚ 0.192) | 0.65 (23.47‚ 2.05) | 12.65 (11.47‚ 14.05) |
| GATA3 | GSE113883 | 3.13 (2.83‚ 3.46) | 0.101 (0.051‚ 0.169) | 0.43 (21.56‚ 2.81) | 12.43 (9.56‚ 14.81) |
| GATA3 | GSE223613 | 4.62 (4.49‚ 4.75) | 0.122 (0.070‚ 0.190) | 22.91 (21.18‚ 0.13) | 10.91 (9.18‚ 12.13) |
| GATA3 | GSE39445 | 8.88 (8.78‚ 8.99) | 0.119 (0.078‚ 0.161) | 23.47 (21.63‚ 0.89) | 11.47 (9.63‚ 12.89) |
| IL13RA1 | GSE113883 | 5.17 (4.92‚ 5.42) | 0.244 (0.162‚ 0.339) | 8.62 (7.17‚ 10.34) | 20.62 (19.17‚ 22.34) |
| IL13RA1 | GSE223613 | 7.10 (6.91‚ 7.30) | 0.288 (0.230‚ 0.356) | 11.69 (11.01‚ 12.22) | 23.69 (23.01‚ 0.22) |
| IL13RA1 | GSE39445 | 10.85 (10.76‚ 10.95) | 0.285 (0.241‚ 0.336) | 12.79 (11.85‚ 13.70) | 0.79 (23.85‚ 1.70) |
| IL18 | GSE113883 | 2.68 (2.53‚ 2.85) | 0.112 (0.051‚ 0.186) | 4.82 (1.91‚ 6.26) | 16.82 (13.91‚ 18.26) |
| IL18 | GSE223613 | 1.92 (1.73‚ 2.15) | 0.041 (0.011‚ 0.124) | 18.37 (13.31‚ 2.98) | 6.37 (1.31‚ 14.98) |
| IL18 | GSE39445 | 7.09 (6.96‚ 7.21) | 0.065 (0.027‚ 0.107) | 12.52 (9.71‚ 15.27) | 0.52 (21.71‚ 3.27) |
| IL1B | GSE113883 | 5.59 (5.29‚ 5.90) | 0.203 (0.142‚ 0.291) | 17.82 (16.35‚ 19.52) | 5.82 (4.35‚ 7.52) |
| IL1B | GSE223613 | 5.51 (5.19‚ 5.86) | 0.269 (0.192‚ 0.366) | 15.09 (14.23‚ 16.49) | 3.09 (2.23‚ 4.49) |
| IL1B | GSE39445 | 12.70 (12.55‚ 12.84) | 0.216 (0.166‚ 0.272) | 17.43 (16.47‚ 18.57) | 5.43 (4.47‚ 6.57) |
| IL1RL1 | GSE113883 | 0.57 (0.42‚ 0.76) | 0.102 (0.045‚ 0.171) | 6.67 (4.88‚ 8.80) | 18.67 (16.88‚ 20.80) |
| IL1RL1 | GSE223613 | 3.24 (2.84‚ 3.65) | 0.122 (0.062‚ 0.226) | 10.77 (5.60‚ 13.22) | 22.77 (17.60‚ 1.22) |
| IL1RL1 | GSE39445 | 3.56 (3.31, 3.83) | 0.163 (0.074, 0.284) | 14.97 (12.11, 18.26) | 2.97 (0.11, 6.26) |
| IL4R | GSE113883 | 6.87 (6.67‚ 7.05) | 0.111 (0.058‚ 0.188) | 0.21 (20.49‚ 2.63) | 12.21 (8.49‚ 14.63) |
| IL4R | GSE223613 | 7.86 (7.63‚ 8.08) | 0.042 (0.010‚ 0.093) | 1.61 (21.79‚ 9.23) | 13.61 (9.79‚ 21.23) |
| IL4R | GSE39445 | 14.74 (14.60‚ 14.90) | 0.093 (0.059‚ 0.140) | 22.46 (19.99‚ 0.42) | 10.46 (7.99‚ 12.42) |
| IL5RA | GSE113883 | 2.68 (2.26‚ 3.20) | 0.274 (0.140‚ 0.415) | 6.11 (4.60‚ 8.13) | 18.11 (16.60‚ 20.13) |
| IL5RA | GSE223613 | 4.82 (4.38‚ 5.26) | 0.126 (0.046‚ 0.228) | 6.57 (4.11‚ 9.72) | 18.57 (16.11‚ 21.72) |
| IL5RA | GSE39445 | 6.07 (5.88‚ 6.26) | 0.114 (0.073‚ 0.160) | 12.07 (9.95‚ 14.33) | 0.07 (21.95‚ 2.33) |
| KIT | GSE113883 | 0.51 (0.41‚ 0.63) | 0.106 (0.070‚ 0.144) | 8.58 (7.49‚ 10.02) | 20.58 (19.49‚ 22.02) |
| KIT | GSE223613 | 1.94 (1.60‚ 2.22) | 0.162 (0.095‚ 0.240) | 12.04 (10.31‚ 14.05) | 0.04 (22.31‚ 2.05) |
| KIT | GSE39445 | 5.80 (5.61, 5.98) | 0.217 (0.147, 0.298) | 12.39 (11.15, 13.52) | 0.39 (23.15, 1.52) |
| MMP9 | GSE113883 | 5.57 (5.06‚ 6.03) | 0.428 (0.254‚ 0.680) | 13.26 (10.65‚ 14.90) | 1.26 (22.65‚ 2.90) |
| MMP9 | GSE223613 | 6.58 (6.31‚ 6.85) | 0.328 (0.156‚ 0.503) | 13.29 (11.87‚ 15.28) | 1.29 (23.87‚ 3.28) |
| MMP9 | GSE39445 | 13.70 (13.39‚ 13.96) | 0.278 (0.173‚ 0.394) | 14.03 (12.10‚ 15.63) | 2.03 (0.10‚ 3.63) |
| SIGLEC8 | GSE113883 | 2.55 (2.04‚ 3.19) | 0.143 (0.043‚ 0.279) | 4.06 (0.57‚ 8.09) | 16.06 (12.57‚ 20.09) |
| SIGLEC8 | GSE223613 | 3.62 (3.07‚ 4.20) | 0.194 (0.024‚ 0.442) | 3.32 (0.33‚ 6.87) | 15.32 (12.33‚ 18.87) |
| SIGLEC8 | GSE39445 | 6.18 (5.90‚ 6.44) | 0.116 (0.052‚ 0.190) | 1.76 (23.83‚ 3.74) | 13.76 (11.83‚ 15.74) |
| STAT6 | GSE113883 | 8.34 (8.18‚ 8.50) | 0.097 (0.030‚ 0.179) | 13.64 (10.88‚ 16.34) | 1.64 (22.88‚ 4.34) |
| STAT6 | GSE223613 | 9.30 (9.20‚ 9.38) | 0.097 (0.054‚ 0.137) | 13.45 (12.56‚ 14.68) | 1.45 (0.56‚ 2.68) |
| STAT6 | GSE39445 | 14.37 (14.01‚ 14.70) | 0.075 (0.032‚ 0.125) | 15.19 (12.76‚ 17.72) | 3.19 (0.76‚ 5.72) |
| TGFB1 | GSE113883 | 7.63 (7.41‚ 7.86) | 0.259 (0.197‚ 0.337) | 13.25 (12.15‚ 14.69) | 1.25 (0.15‚ 2.69) |
| TGFB1 | GSE223613 | 8.85 (8.79‚ 8.92) | 0.095 (0.054‚ 0.135) | 12.68 (11.65‚ 13.80) | 0.68 (23.65‚ 1.80) |
| TGFB1 | GSE39445 | 10.94 (10.83‚ 11.04) | 0.169 (0.125‚ 0.219) | 13.35 (11.93‚ 14.57) | 1.35 (23.93‚ 2.57) |
| TIMP1 | GSE113883 | 7.88 (7.60‚ 8.15) | 0.316 (0.232‚ 0.402) | 14.07 (13.15‚ 15.04) | 2.07 (1.15‚ 3.04) |
| TIMP1 | GSE223613 | 4.73 (4.60‚ 4.88) | 0.100 (0.054‚ 0.151) | 14.89 (13.46‚ 16.48) | 2.89 (1.46‚ 4.48) |
| TIMP1 | GSE39445 | 11.10 (10.98‚ 11.22) | 0.121 (0.083‚ 0.166) | 14.13 (12.53‚ 15.66) | 2.13 (0.53‚ 3.66) |
| TNFSF4 | GSE113883 | 0.91 (0.76‚ 1.06) | 0.058 (0.012‚ 0.103) | 5.77 (3.22‚ 7.76) | 17.77 (15.22‚ 19.76) |
| TNFSF4 | GSE223613 | 4.26 (4.02‚ 4.46) | 0.136 (0.066‚ 0.217) | 14.28 (12.43‚ 15.36) | 2.28 (0.43‚ 3.36) |
| TNFSF4 | GSE39445 | 7.31 (7.14‚ 7.48) | 0.178 (0.118‚ 0.245) | 14.33 (13.14‚ 15.84) | 2.33 (1.14‚ 3.84) |
| GSE113883 used log(TPM + 1); GSE22613 TMM-normalized log2-CPM; and GSE39445, log2 quantile normalized microarray intensity | | | | | |
| CI, Confidence Intervals; MESOR- Midline Estimating Statistic of Rhythm  *Amplitude is the difference between the fitted MESOR and peak.  ^#^A one-unit difference on the log2 scale corresponds to a doubling of the underlying quantity. | | | | | |

| **Table S3**: Evaluation of known in-phase and anti-phase clock gene relationships \| \| \| \| \| \| \|; Clock gene pair \| Dataset \| Expected relationship \| Circular difference in phase with bootstrapped 95% CI \| \| Zero-lag Pearson cross correlation coefficient \| \|; *BMAL‚ REV-ERBα* \| GSE223613 \| Antiphase \| 10.51 (9.64‚ 11.55) \| \| -0.881 \| \|; *BMAL‚ REV-ERBα* \| GSE39445 \| Antiphase \| 11.55 (6.66‚ 11.95) \| \| -0.627 \| \|; *BMAL‚ REV-ERBβ* \| GSE223613 \| Antiphase \| 11.11 (10.28‚ 11.93) \| \| -0.932 \| \|; *BMAL‚ REV-ERBβ* \| GSE39445 \| Antiphase \| 11.20 (6.82‚ 11.95) \| \| -0.708 \| \|; *REV-ERBα ‚ REV-ERBβ* \| GSE113883 \| In phase \| 2.30 (0.18‚ 4.86) \| \| +0.496 \| \|; *REV-ERBα ‚ REV-ERBβ* \| GSE223613 \| In phase \| 0.60 (0.19‚ 1.12) \| \| +0.954 \| \|; *REV-ERBα ‚ REV-ERBβ* \| GSE39445 \| In phase \| 1.25 (0.47‚ 2.04) \| \| 0.944 \| \|; *PER1‚ PER2* \| GSE113883 \| In phase \| 2.02 (0.76‚ 3.63) \| \| +0.728 \| \|; *PER1‚ PER2* \| GSE223613 \| In phase \| 1.10 (0.18‚ 2.15) \| \| +0.792 \| \|; *PER1‚ PER2* \| GSE39445 \| In phase \| 1.07 (0.06‚ 2.72) \| \| +0.644 \| \|; *PER1‚ PER3* \| GSE113883 \| In phase \| 2.55 (1.18‚ 3.89) \| \| +0.754 \| \|; *PER1‚ PER3* \| GSE223613 \| In phase \| 0.95 (0.06‚ 2.10) \| \| +0.878 \| \|; *PER1‚ PER3* \| GSE39445 \| In phase \| 0.81 (0.06‚ 1.85) \| \| +0.707 \| \|; *PER2‚ PER3* \| GSE113883 \| In phase \| 0.53 (0.03‚ 1.32) \| \| +0.878 \| \|; *PER2‚ PER3* \| GSE223613 \| In phase \| 2.05 (0.56‚ 3.71) \| \| +0.811 \| \|; *PER2‚ PER3* \| GSE39445 \| In phase \| 0.26 (0.02‚ 1.80) \| \| +0.974 \| \|; *Circular difference is the shortest distance across the 24h clock. \| \| \| \| \| \| \|; Bootstrapped estimates (2,000 iterations) \| \| \| \| \| \| \| **Table S4: Sensitivity Analyses- Leave One Participant Out Analyses in the included Cohorts** | | | |
| --- | --- | --- | --- |
| **Measure** | **GSE113883** | **GSE223613** | **GSE39445** |
| Replicated asthma genes in primary analyses | 23 | 23 | 23 |
| Primarily rhythmic in cohort in primary analyses | 20 | 20 | 21 |
| Significant in every LOPO refit | 19 | 19 | 21 |
| Percentage robustly rhythmic | *95% | *95% | 100% |
| Median maximum acrophase shift (h) | 0.60 | 0.57 | 0.41 |
| Largest maximum acrophase shift (h) | 1.07 | 1.23 | 0.90 |
| Median minimum amplitude ratio | 0.863 | 0.869 | 0.909 |
| Median maximum amplitude ratio | 1.13 | 1.14 | 1.09 |
| Failed model fits | 0 | 0 | 0 |
| Rhythmicity retention is based on nominal p < 0.05 in the LOPO refits, among genes significant at FDR < 0.05 in each cohort's primary analysis. | | | |
| *GSE113883- *STAT6* lost significance; GSE223613- *SIGLEC8* lost significance | | |  |

| **Table S5**: Expression-filter sensitivity results for the bulk RNA sequencing data | | | | |
| --- | --- | --- | --- | --- |
| **Cohort** | **Threshold** | **Evaluable asthma genes** | **# of 23 replicated genes evaluable** | **Replicated genes rhythmic** |
| GSE113883 | TPM ≥0.01 | 33 | 23 | 20 |
|  | TPM ≥0.05 | 33 | 23 | 20 |
|  | TPM ≥0.10 (primary analyses) | 33 | 23 | 20 |
|  | TPM ≥0.50 | 24 | 21 | 18 |
|  | TPM ≥1.00 | 22 | 20 | 18 |
| GSE223613 | CPM ≥0.05 | 33 | 23 | 20 |
|  | CPM ≥0.10 (primary analyses) | 33 | 23 | 20 |
|  | CPM ≥0.50 | 28 | 23 | 20 |
|  | CPM ≥1.00 | 26 | 23 | 20 |

| **Table S6**: Reducing expression-filter threshold to ≥25% of samples (from ≥50% in primary analyses): | | | |
| --- | --- | --- | --- |
| **Cohort** | **%samples that passed threshold** | **Peak phase** | **P-value** |
| GSE223613  *RORB* (clock gene) | 39.2% | 12.10 | 0.088 |
| *IL33* (asthma-related gene) | 38.0% | 11.52 | 0.203 |
